# Contextualizing molecular and structural aging across human organs

**DOI:** 10.1101/2025.05.26.25328375

**Authors:** Juan Shu, Yuxin Guo, Julio Chirinos, Zirui Fan, Yilin Yang, Yujue Li, Xiaochen Yang, Cameron Beeche, Buxin Su, Iris Horng, Rong Zheng, Peristera Paschou, Li-San Wang, Joan M. O’Brien, Ruben Gur, Jinjie Lin, Walter Witschey, Daniel J. Rader, Anthony Rosenzweig, Bingxin Zhao

## Abstract

Organ-specific aging clocks have shown promise as predictors of disease risk and aging trajectories; however, the underlying biological mechanisms they reflect remain largely unexplored. Here, we use large-scale proteomic and imaging data to investigate the relationships among organ-specific and modality-specific aging clocks and to uncover the biological processes they represent. By estimating paired protein-based and imaging-based aging clocks across 8 major organs, we demonstrate that these omics and structural profiles exhibit distinct phenotypic and genetic signatures, each potentially quantifying different stages and playing complementary roles within a unified biological aging process. Furthermore, context-specific aging clocks from multiple organs often converge and jointly capture established biological and disease pathways. For example, 65.7% of the KEGG Alzheimer’s disease pathway is enriched by at least one of 11 protein- and imaging-based aging clocks, with each clock representing different components of the pathway. These results underscore the importance of a pan-organ multi-modal perspective for quantifying the mechanisms underlying age-related diseases. Additionally, we identify modality-specific links between aging clocks and complex diseases and lifestyle factors. In summary, we uncover intricate relationships among molecular and structural aging clocks across human organs, providing novel insights into their context-specific roles in capturing consequences of aging biology and their implications for disease risk.

---

Aging is a fundamental biological process characterized by an eventual gradual decline in organ function and a series of prominent hallmarks^1,2^. Aging represents a critical risk factor for chronic diseases and their multimorbidity^3–5^ spanning cardiovascular diseases^6^, brain disorders^7^, and metabolic conditions^8^. Although aging is universal, its underlying mechanisms are complex and vary widely across organs and between individuals^9–12^, involving specific molecular changes and structural deteriorations, shaped by the interplay of genetic and environmental factors^13^. Since the direct assessment of these complex mechanisms in humans is challenging, organ-specific aging clocks using predictive aging biomarkers, including proteomic^14–17^ and imaging^18,19^ data, have been recently developed to estimate biological aging, aiming to capture the downstream consequences of underlying factors that promote or mitigate aging^20^. The accessibility of both proteomic and imaging data is expected to increase substantially in global biobanks. For example, the UK Biobank (UKB) study plans to generate proteomic data for half a million participants and imaging data for nearly 100,000 participants^21^. Given these developments, understanding context-specific biological aging patterns in large-scale cohorts through protein-based and imaging-based aging clocks as surrogate markers can play a crucial role in aging research. Such knowledge can guide the design of targeted interventions and preventive strategies to promote healthy aging and reduce disease risk in precision medicine.

Despite strong evidence independently supporting proteomic and imaging data as promising indicators and effective tools for quantifying aging trajectories, the relationships among these molecular and structural aging clocks across human organs remain unclear. Moreover, the biological mechanisms reflected by these organ-specific and modality-specific aging clocks are still largely unexplored. Notably, due to the inherent differences across data modalities, individual aging clocks may capture only distinct and partial aspects of the complex aging process underlying physiology and disease development. For example, proteomic biomarkers provide molecular and cellular insights by tracking shifts in protein expression that reflect overall^22^ or organ-specific^14–16^ biological aging. These omics aging measures can rapidly respond to molecular changes or inflammation but may fail to fully capture the slow, progressive nature of structural aging within specific organs. Imaging data, such as magnetic resonance imaging (MRI), captures structural, functional, or physiological aging changes at the organ or tissue level^18,19,23–26^, which ultimately reflect underlying molecular processes. Therefore, proteomic and imaging data may represent complementary views of a unified biological aging process: imaging reveals downstream, system-level consequences, while plasma proteomics captures circulating signals that may reflect upstream molecular activity. Understanding the specific molecular or structural alterations captured by each aging clock, and how they relate to one another, is essential for gaining deeper insights into their implications for disease mechanisms and risk prevention.

Here we present a pan-organ analysis to explore the interactions of molecular and structural aging clocks and the underlying biological mechanisms they reflect. Using large-scale multi-modal data from the UKB study, a well-characterized cohort of relatively healthy adults, we constructed 10 protein-based and 14 imaging-based aging clocks, including paired clocks for 8 major organs. We systematically characterized cohort- and individual-level connections and differences of these organ-specific and modality-specific aging clocks. We further uncovered their distinct genetic architecture^27^, along with the involved gene modules in network biology, and their complementary roles in established biological and disease pathways^28,29^. Furthermore, we linked these context-specific aging clocks to age-related diseases and lifestyle factors, identifying their genetic and phenotypic associations, as well as putative causal relationships. An overview of the study design is illustrated in **Figure 1**.

**Fig. 1.**
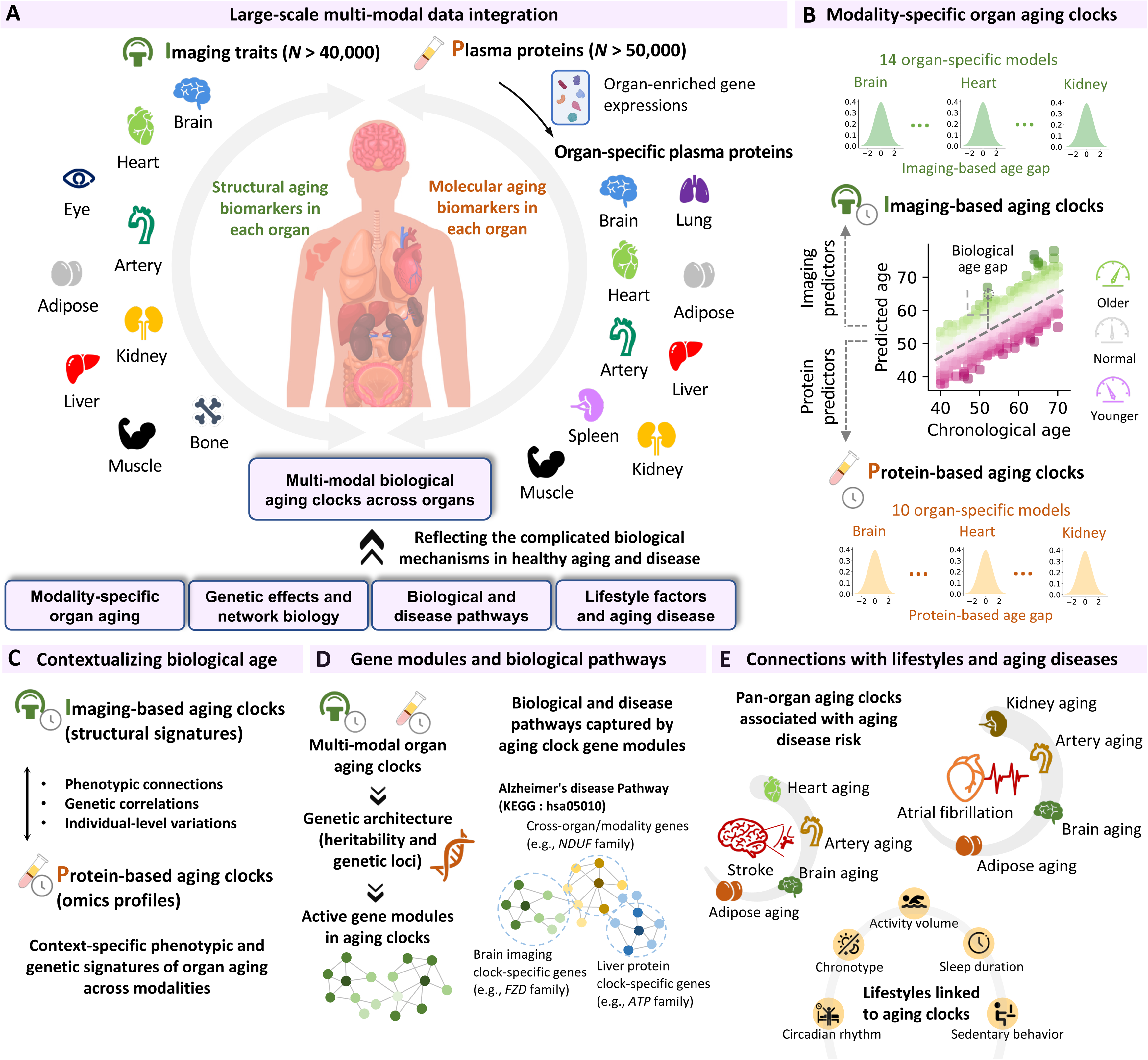
Overview of study design and analyses. **(A).** An overview of our multi-modal organ-specific biological aging study, which evaluates modality-specific aging clocks across a broad range of organs (and tissues) using imaging and proteomic data to capture structural and molecular aspects of aging, respectively. (**B**) An illustration of the model used to derive organ-specific and modality-specific aging clocks. We used the derived aging clocks to contextualize biological age by examining the relationships among these clocks (**C**), uncovering the underlying biological pathways they represent through network biology analysis (**D**), and evaluating their associations with lifestyle factors and complex diseases (**E**).

## RESULTS

### Pan-organ molecular and structural aging clocks

We developed modality-specific aging clocks separately using imaging and plasma protein data across a broad range of organs (**Methods**; average sample size *n* = 54,150 for imaging and 44,803 for proteins). Following previous studies^14–16^, we defined plasma proteins as organ-specific if their gene expression in a particular organ was at least four times higher than in any other organ^30^, and used these proteins to construct organ-specific protein-based aging clocks. In total, we developed 14 imaging-based aging clocks and 10 protein-based aging clocks and assessed their pairwise phenotypic associations. We then performed genome-wide association studies (GWAS) (**Methods**) and estimated their pairwise genetic correlations using LDSC^31^. The detailed genetic architecture results are provided in a later section. Below, we characterized the cohort-level features of these aging clocks, focusing on organ-wise connections within each modality as well as cross-modality specificities.

**Figure 2A** illustrates the phenotypic (upper triangle above the diagonal line) and genetic (lower triangle under the diagonal line) correlations among imaging-based aging clocks across various organs (and tissues). We found that these organ-specific aging clocks primarily captured distinct aging dynamics within each organ. Nevertheless, several notable clusters of phenotypically correlated clock pairs emerged, indicating a moderate to low degree of inter-organ aging connections among several organs (false discovery rate [FDR] multiple testing-adjusted, β range = [-0.03, 0.56], *P* < 6.94×10^-5^). Furthermore, a subset of these phenotypic links were supported by underlying genetic correlations, with genetic correlation coefficients consistently larger than the corresponding phenotypic correlations (β range = [-0.3, 0.58], *P* < 6.04×10^-5^, **Fig. 2B**). In particular, brain aging clocks shared moderate genetic similarities across structural MRI (sMRI), diffusion MRI (dMRI), and resting-state functional MRI (rfMRI) modalities (β range = [0.53, 0.62], *P* < 9.23×10^-23^), and had positive genetic associations with the bone composition aging clock (β range = [0.24, 0.28], *P* < 3.62×10^-5^). These results might reflect potential shared aging mechanisms such as inflammation and mitochondrial dysfunction^32^, as well as the brain-bone axis mediated by hypothalamic leptin signaling^33^. Additionally, the adipose, muscle, and body composition aging clocks had genetic correlations with those of the heart, kidney, and liver (β range = [0.30, 0.58], *P* < 5.55×10^-5^). These inter-organ connections may indicate shared systemic mechanisms such as lipotoxicity^34^, oxidative stress^35^, and mitochondrial dysfunction^36^. We also observed genetic connections between the heart and artery aging clocks (β = 0.25, *P* < 2×10^-5^), consistent with aging across the cardiovascular system^37^. Furthermore, we identified several significant but relatively weak phenotypic connections, for example, brain aging clocks were phenotypically correlated with those of the heart, arteries, adipose tissue, muscle, and body composition (β range = [0.04, 0.10], *P* < 1.1×10^-6^). Although global genetic correlations between these aging clocks were not statistically significant, these weak phenotypic associations likely suggest potential shared influences, possibly reflecting overlapping biological pathways or localized genetic effects not detected at the genome-wide level.

**Fig. 2.**
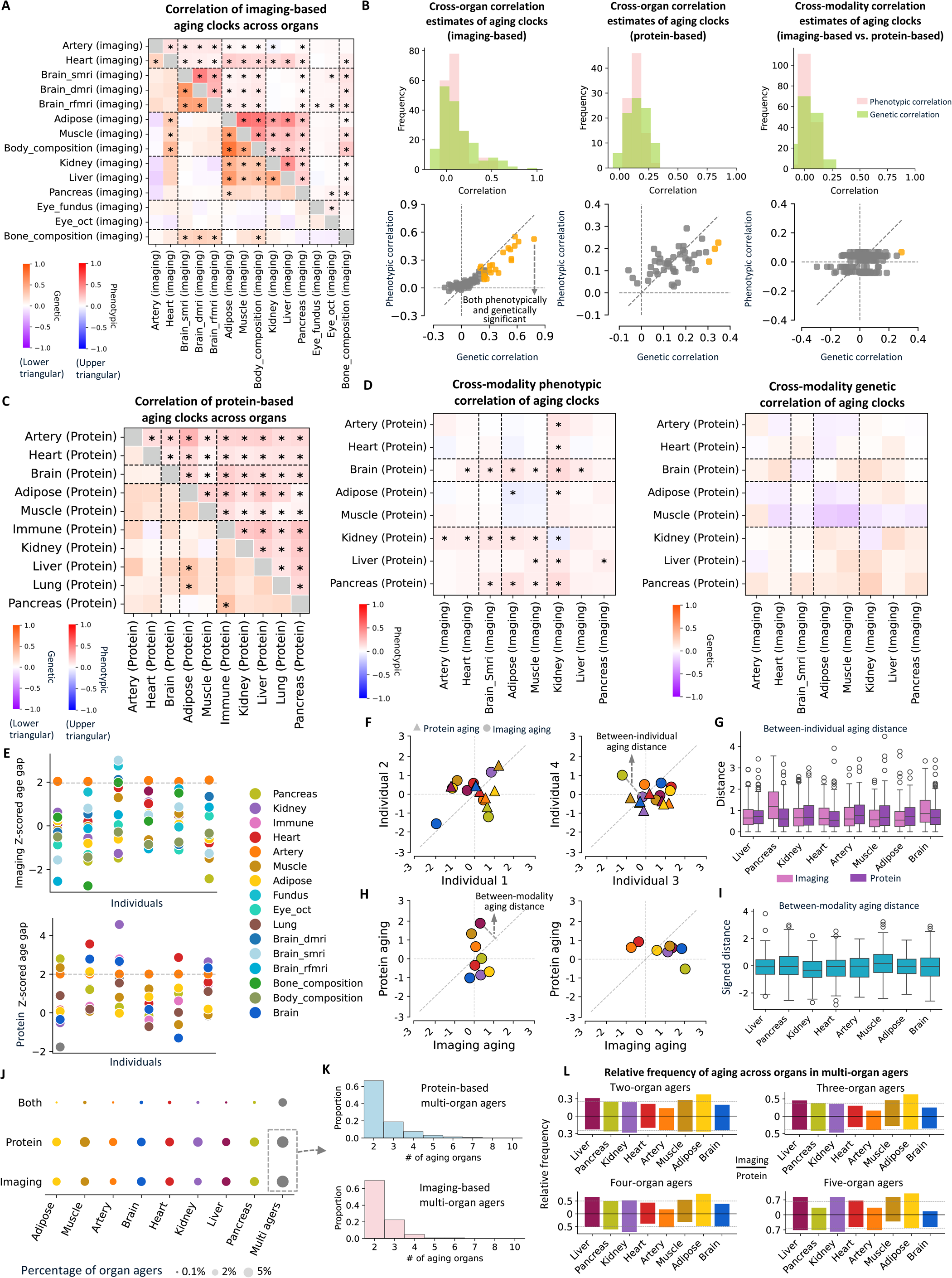
Contextualizing aging clocks. (A). Phenotypic (upper triangle above the diagonal line) and genetic (lower triangle under the diagonal line) correlations among imaging-based aging clocks. Asterisks indicate pairs with false discovery rate (FDR)-adjusted *P*-values < 0.05. (**B**). The top panel shows the distribution of genetic and phenotypic correlations across three categories: within imaging-based clocks (left), within protein-based clocks (middle), and between modalities (right). The bottom panel compares genetic and phenotypic correlations for each category. Orange indicates pairs with both significant genetic and phenotypic correlations. (**C**). Phenotypic (upper triangle) and genetic (lower triangle) correlations among protein-based aging clocks, with asterisks indicating FDR-adjusted *P*-values < 0.05. (**D**). Phenotypic (left) and genetic (right) correlations between imaging-based and protein-based aging clocks. Asterisks indicate correlations with FDR-adjusted *P*-values < 0.05. (**E**). Individuals with similar aging scores for a given organ (e.g., an artery aging clock score of 2) often displayed distinct aging patterns across other organs within the same modality, shown in the top panel for imaging-based clocks and the bottom panel for protein-based clocks. Six participants were randomly selected and are presented as examples. (**F**). Examples of aging clock variations between a pair of individuals: some aging clocks show similar values, while others exhibit larger differences. (**G**). Between-individual pairwise Euclidean distances across all individual pairs, calculated separately for each organ and modality. (**H**). Examples of aging clock variations within a single individual for paired organ aging clocks, showing that imaging-based and protein-based clocks for the same organ can yield different scores. (**I**). Between-modality Euclidean distances across all individuals, calculated separately for each organ. (**J**). Proportion of individuals classified as organ-specific or multi-organ agers based on imaging-based aging clocks, protein-based aging clocks, or both modalities. (**K**). Distribution of the number of aging organs among multi-organ agers, shown separately for protein-based (top) and imaging-based (bottom) aging clocks. (**L**). Organ-specific aging frequency among multi-organ agers with aging observed in k organs (k = 2, 3, 4, or 5). Each subfigure includes a reference line at k/10 (i.e., 0.2, 0.3, 0.4, or 0.5), representing the situation of equal contribution across the 10 organs analyzed in the multi-organ aging assessment.

We also evaluated the inter-organ correlations of protein-based aging clocks (**Fig. 2C**). While aging clocks from all organs showed statistically significant phenotypic correlations, most were weak (β range= [0.02, 0.29], *P* < 6.95×10^-10^). These weak correlations are consistent with previous studies emphasizing organ-specificity of protein-based aging clocks across diverse cohorts^14–16^. Consistent with the weak phenotypic correlations, genetic correlations among protein-based aging clocks were also limited, with only a few pairs reaching statistical significance and mostly showing small effect sizes (β range= [-0.06, 0.35], *P* < 3×10^-4^). As a result, the alignment between phenotypic and genetic correlation coefficients was weaker for protein-based aging clocks compared to imaging-based ones (**Fig. 2B**). These findings indicate that, although protein expression levels are strongly influenced by underlying genetic variations^38^, cohort-level phenotypic and genetic correlations among organ-specific aging clocks are relatively weak. This underscores their organ-specific nature and suggests that they may capture context-dependent aging processes.

Next, we examined the cross-modality correlations between paired protein-based and imaging-based aging clocks across 8 organs (**Fig. 2D**). Among within-organ aging clock pairs, only the brain and immune system exhibited significant positive phenotypic correlations between their protein-based and imaging-based clocks (β range = [0.06, 0.08], *P* < 8.27×10^-5^). In contrast, adipose and kidney showed negative phenotypic correlations (β range = [-0.08, -0.06], *P* < 3.19×10^-4^). For all other organs, protein-based and imaging-based clocks were largely independent, suggesting they captured distinct aspects of biological aging. Regarding cross-organ pairs, most significant pairs showed weak positive correlations (β range = [-0.09, 0.15], *P* < 2.78×10^-4^). For example, the brain protein-based aging clock correlated with imaging-based aging clocks of the brain, heart, adipose, and abdominal organs (spleen, kidney, liver, β range = [0.05, 0.14], *P* < 9.06×10^-5^). Additionally, both protein-based and imaging-based kidney aging clocks showed cross-modality associations with multiple other organs, consistent with the kidney’s central role in systemic aging^39,40^. In contrast to phenotypic correlation, only one significant cross-modality genetic correlation was observed between modality-specific aging clocks (**Fig. 2B**), which is between imaging-based aging clock of pancreas and protein-based aging clock of spleen (β = 0.29, *P* = 1×10^-4^). Overall, these results suggest that, while some notable links exist, organ-specific protein-based and imaging-based aging clocks may differ substantially, providing complementary insights into context-specific aging processes.

### Quantifying individual-level aging variations

The generally weak correlations and relative independence among aging clocks at the cohort level suggest that, at the individual level, aging in one organ is not necessarily linked to aging in others, and within a single organ, protein-based and imaging-based aging clocks may also diverge. We conducted additional analyses to further illustrate these individual-level variations. Within both protein-based and imaging-based aging clocks, individuals with similar aging scores for one organ often exhibited varying aging patterns across other organs **(Fig. 2E**). This complexity increased when both modalities were considered together to compare aging clocks between individuals (**Fig. 2F**). To quantify this variability, we calculated between-individual differences in estimated aging clocks within each organ and modality. Certain clocks, such as the imaging-based aging clocks for the brain and pancreas, exhibited greater between-individual distances and higher variability (**Fig. 2G**). Additionally, for a given individual and organ, it is possible to have a high aging score in imaging-based clock but a low score in protein-based clock, or vice versa (**Fig. 2H**). The distribution of such between-modality aging distance showed a consistent pattern across all organs (**Fig. 2I**).

We further examined extreme agers across modalities, defined as individuals with normalized aging scores greater than two^14^. Both imaging-based and protein-based aging clocks indicated that approximately 2% of individuals qualified as extreme agers in each organ, while around 5% were identified as multi-organ agers, those experiencing aging in at least two organs (**Fig. 2J**). Notably, fewer than 0.3% of individuals exhibited organ aging detected by both modalities within the same organ. However, this proportion increased to 2.6% among multi-organ agers, who may experience more systemic and advanced biological aging, encompassing both molecular (protein-based) and structural (imaging-based) changes. Among multi-organ agers, most individuals exhibited aging in two or three organs, with protein-based aging clocks showing a slightly higher proportion of individuals aging in more than three organs (**Fig. 2K**). Additionally, some organs appeared more frequently in individuals with multi-organ aging, particularly abdominal organs such as the liver, kidney, pancreas, and spleen, as well as adipose (**Fig. 2L**). These organs serve as metabolic and detoxification hubs essential for maintaining whole-body homeostasis. Their functions, ranging from filtering waste and regulating intermediary metabolism, to promoting immune responses, make them especially sensitive to systemic stress and cumulative physiological burden over time. As a result, they may show signs of aging earlier and more consistently across individuals with widespread physiological decline^1,41,42^.

### Context-specific and pleiotropic gene modules underlying aging clocks

We conducted GWAS using a half-half random split discovery-replication design to identify and replicate genetic variants associated with modality-specific aging clocks (**Methods**). At a stringent threshold of *P* < 2.08×10^-9^ (5×10^-8^/24, the number of aging clocks analyzed), our discovery GWAS identified 1,336 independent (Linkage disequilibrium [LD] *r*^2^ < 0.1) significant associations^43^ (**Table S1**). The majority of these associations, 94% for protein-based and 88% for imaging-based aging clocks, met the replication threshold (*P* < 0.05/1,336) in the independent replication GWAS. They also demonstrated strong consistency in genetic effect estimates (**Figs. S1-S2**). Applying a stricter criterion that both discovery and replication GWAS met the genome-wide significance threshold (*P* < 2.08×10^-9^), we identified 913 independent significant associations for protein-based aging clocks and 59 for imaging-based aging clocks, corresponding to 68 unique independent organ-cytoband pairs for protein-based aging clocks and 20 for imaging-based aging clocks (**Fig. 3A**). Additionally, the mean heritability^44^ was 0.383 (*h*^2^ range = [0.209, 0.572], *P* < 4.90×10^-9^) for protein-based aging clocks and 0.377 (*h*^2^ range = [0.194, 0.539], *P* < 1.19×10^-15^) for imaging-based aging clocks (**Table S2**). These results suggest that both imaging-based and protein-based aging clocks were heritable, yet their identified genetic loci were largely non-overlapping. These findings support the presence of distinct genetic architectures underlying modality-specific aging clocks, in line with moderate genetic correlation presented in earlier sections.

**Fig. 3.**
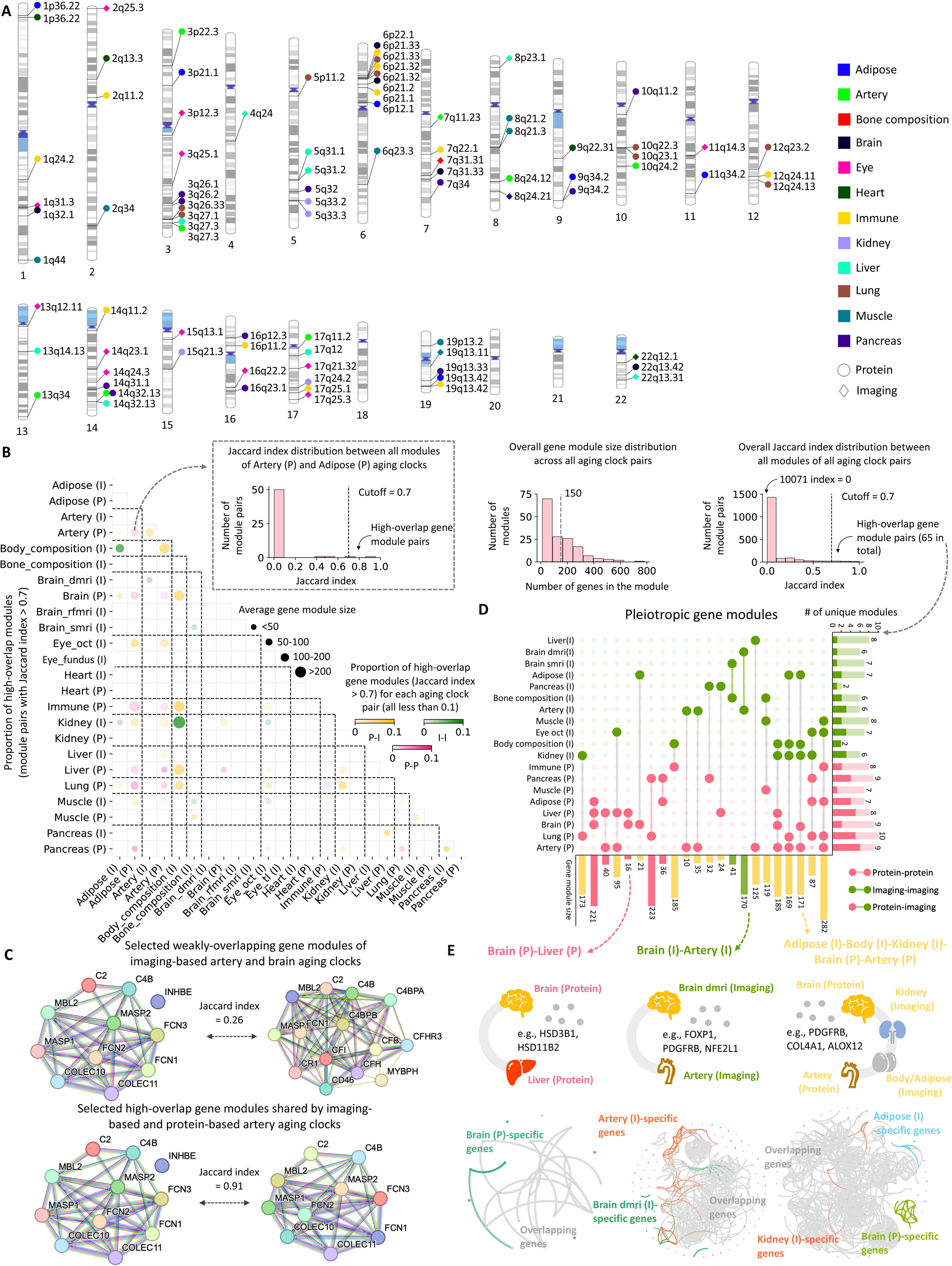
Genetic architecture and gene modules. (**A**). Ideogram of genomic regions associated with protein-based and imaging-based aging clocks (*P* < 2.08×10^-9^). (**B**). The left triangle panel shows the proportion of high-overlap gene modules (Jaccard index > 0.7) for each pair of aging clocks. As an example, the top left histogram provides a zoomed-in view of the Jaccard index distribution for all gene modules from protein-based aging clocks of the artery and adipose. In the legend, P-I denotes organ pairs between protein-based and imaging-based aging clocks, I-I refers to pairs from imaging-based clocks, and P-P indicates pairs from protein-based clocks. The top middle histogram displays the distribution of module sizes, while the top right histogram shows the overall distribution of Jaccard indices. (**C**). Example of weakly-overlapping gene modules (top panel) and high-overlap gene modules (bottom panel). (**D**) Visualizing the 22 pleiotropic gene modules. Each row represents an organ-specific aging clock, either imaging-based (I) or protein-based (P), along with the number of associated gene modules. Each column corresponds to a pleiotropic gene module, indicating the aging clocks involved and the module size. (**E**). Three zoomed-in examples of pleiotropic gene modules in (**D**) are shown. The top panels highlight the aging clocks involved and key genes within each pleiotropic module, while the bottom panels show the view of corresponding protein-protein interaction subnetworks, with annotations distinguishing aging clock-specific genes and pleiotropic genes (gray).

The observed differences in genetic architecture suggest that context-specific aging clocks may represent divergent aging mechanisms in downstream genomic functions and biological processes. It is well-established that genes involved in similar functions or diseases tend to interact within networks rather than act in isolation^45,46^. To gain insights into their functional roles, we performed a network biology analysis on these genetic signals. Briefly, we integrated aging clock-associated genes with external protein-protein interaction databases^47–51^, applied network propagation to form interacting gene modules^52^, and identified active gene modules for each aging clock (**Methods**). Overall, we identified 80 gene modules for 10 protein-based aging clocks and 78 gene modules for 14 imaging-based aging clocks with an average gene module size of 150 (**Tables S3-S4**).

To evaluate the functional similarity among aging clocks across different modalities and organs, we quantified the overlap pattern of their gene modules using the Jaccard index. Among all gene module pairs, only 15.3% exhibited any overlap (Jaccard index > 0), and a mere 65 pairs (0.5%) had high overlap (Jaccard index > 0.7, **Fig. 3B**). Representative examples of these gene modules are shown in **Figure 3C**. Across all aging clock pairs, fewer than 10% of their corresponding gene module pairs exhibited high overlap (Jaccard index > 0.7), suggesting that the vast majority (over 90%) of gene modules associated with aging clocks were distinct and context-specific in the network biology landscape. Notably, even protein-based and imaging-based aging clocks derived from the same organ showed minimal gene module overlap. For example, only paired clocks from the artery, muscle, and pancreas displayed a few gene module pairs with a Jaccard index exceeding 0.7 (**Fig. 3B**).

While most of the gene modules were distinct, several notable clusters emerged among the 65 pairs of high-overlap gene modules. For example, protein-based aging clocks for the adipose, lung, and artery displayed broader similarity, sharing gene modules with at least 9 other aging clocks across modalities and organs, indicating presence of shared, potentially universal aging programs related to these gene modules. To explore the detailed patterns of these high-overlap gene modules, we reorganized them into an interaction network and identified cliques (**Methods**), resulting in the formation of 22 pleiotropic gene modules (**Fig. 3D**). These pleiotropic modules may span aging clocks from different organs within the same modality (such as protein-based aging clocks of the brain and liver, imaging-based clocks of the brain and artery), as well as more complex cross-modality overlaps that extended across multiple organs and different data modalities (**Fig. 3E**). Different aging clocks within the same pleiotropic gene module shared the majority of genes, while also containing a smaller subset of clock-specific genes unique to each aging clock. For example, we found shared genes of *HSD3B1* and *HSD11B2* in the pleiotropy gene module shared by protein-based aging clocks of the liver and brain, where both genes influence liver function and brain health through systemic hormonal signaling. Specifically, *HSD3B1* contributes to hepatic steroidogenesis and neurosteroid production, while *HSD11B2* protects the brain and liver from glucocorticoid excess by inactivating cortisol, maintaining metabolic and neural homeostasis^53,54^. Additionally, in pleiotropic gene modules involving imaging-based aging clocks of brain and artery, we found several shared genes such as *FOXP1*, *PDGFRB*, and *NFE2L1*, which jointly contribute to brain and arterial health by regulating neurodevelopment, vascular integrity, and oxidative stress responses, coordinating neuronal differentiation, blood-brain barrier stability, and redox homeostasis^55–57^. These results help uncover shared molecular mechanisms and involved genes that drive aging across multiple organs, providing insights into systemic aging processes. Additionally, the gene module-based network approach provides a systematic way to identify the biological processes and disease pathways reflected by aging clocks, which will be further explored in the next section.

### Biological and disease pathways captured by pan-organ aging clocks

Using functional enrichment analysis^58^ (**Methods**), we identified KEGG biological and disease pathways in which the gene modules of aging clocks were significantly enriched after multiple testing adjustment (*P* < 3.6×10^-2^, **Table S5**). Below, we aimed to uncover patterns from three important perspectives. First, we examined results by data modality, highlighting key functional similarities and differences between molecular and structural aging clocks, generated by protein-based and imaging-based data, respectively (**Figs. 4A-4B**). Second, we explored organ-wide specificity within each modality, identifying pathways that are exclusively captured by specific organs, such as pathways uniquely detected by the brain aging clock among all protein-based aging clocks (**Figs. 4C-4D**). Third, we analyzed pathways that were frequently enriched across multiple aging clocks to gain more insights into the biological significance and interpretations of such overlaps (**Figs. 4E-4F**).

**Fig. 4.**
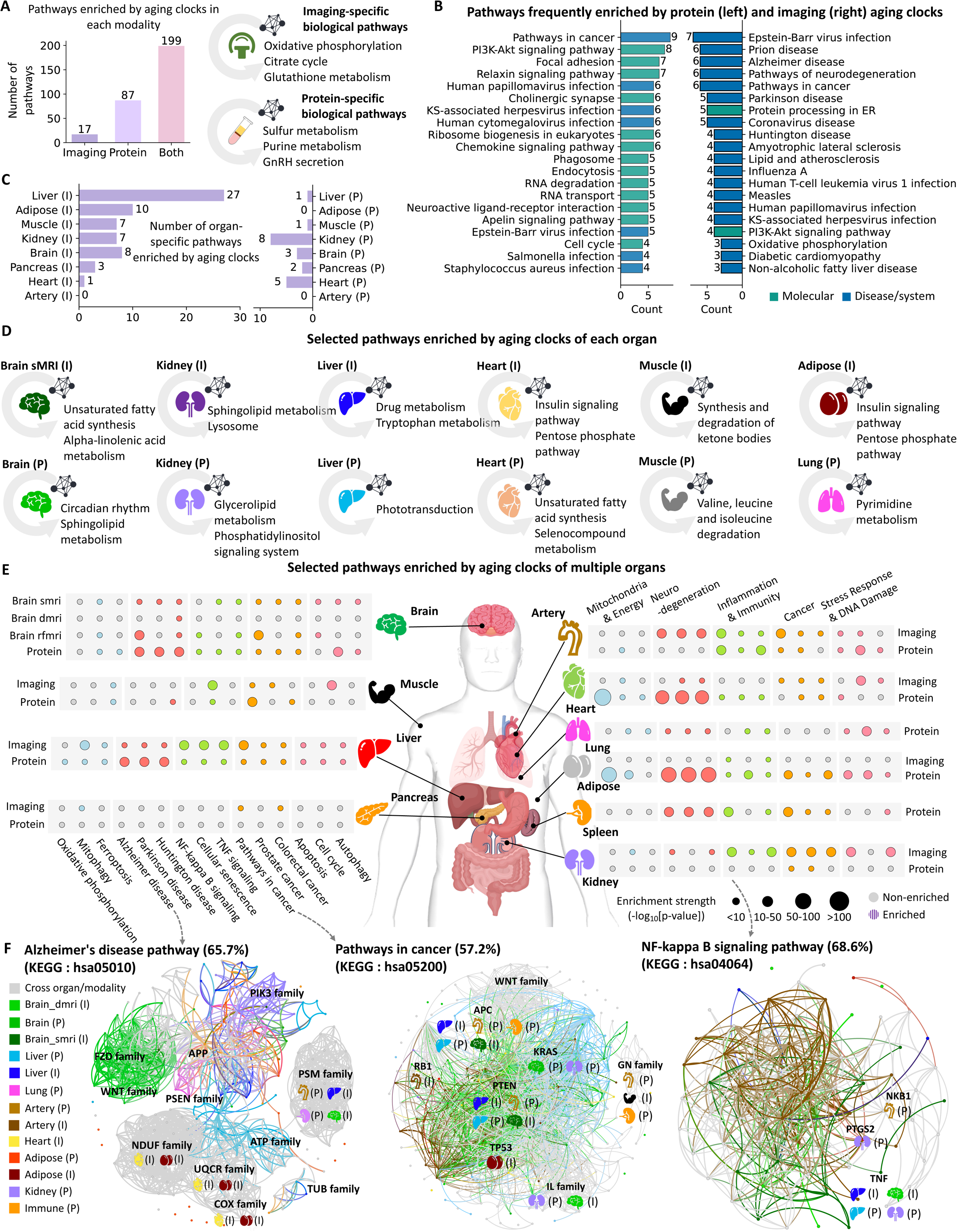
Pathway enrichment. (**A**). The number of KEGG pathways uniquely enriched in imaging-based or protein-based aging clocks, as well as those shared by both. Representative examples of imaging-specific and protein-specific pathways are also provided. (**B**). The top 20 most frequent and strongly enriched pathways (*P* < 1×10^-8^) among protein-based and imaging-based aging clocks. Colors represent either molecular-level pathways or system/disease-level pathways. (**C**). The number of organ-specific enriched pathways identified within imaging-based (left) and protein-based (right) aging clocks, respectively. (**D**). Selected pathways enriched by aging clocks for each organ. The top panels display examples of pathways uniquely enriched by the imaging-based aging clock from a single organ, rather than across multiple organs. The bottom panels illustrate similar organ-specific enrichment patterns observed in protein-based aging clocks. (**E**). Selected pathways frequently enriched across aging clocks from multiple organs, grouped into six categories: mitochondrial and energy, neurodegeneration, inflammation and immunity, cancer, as well as stress response and DNA repair. Each category includes three representative pathways. (**F**). Three zoomed-in examples of enriched pathways from **(E)** are shown. Each example presents the corresponding protein-protein interaction subnetwork, with annotations indicating genes uniquely captured by protein-based (P) or imaging-based (I) aging clocks, as well as genes shared across multiple aging clocks (gray).

There were 17 pathways exclusively associated with imaging-based aging clocks, involving oxidative phosphorylation, the citrate cycle, and glutathione metabolism (**Fig. 4A**). These pathways reflect core metabolic and oxidative stress processes that manifest as structural and functional changes at the organ level^1^. On the other hand, 87 pathways were specific to protein-based aging clocks, such as sulfur metabolism, purine metabolism, and GnRH secretion, which reflect systemic endocrine and metabolic regulation readily captured in plasma proteomics^59^. In addition, a total of 199 pathways were enriched by aging clocks of both two modalities. The greater number of modality-specific pathways in protein-based clocks may relate to the fact that plasma proteins directly reflect active biological processes and systemic physiological changes, capturing diverse molecular signals^59^. To further evaluate this hypothesis, we examined the top 20 most frequent and strongly enriched pathways (*P* < 1×10^-8^) across the two modalities (**Fig. 4B**). We found that protein-based aging clocks predominantly shared molecular-level pathways (13/20), whereas imaging-based aging clocks were more enriched for system- or disease-level pathways (18/20).

We provided more detailed modality-specific comparisons at the organ level. For example, in the heart, protein-based and imaging-based aging clocks were enriched in 122 and 113 pathways, respectively, with 65 pathways overlapping between the two modalities. Protein-specific pathways included protein digestion (*P* = 7.72×10^-90^), aminoacyl-tRNA biosynthesis (*P* = 1.59×10^-32^), and ECM-receptor interaction (*P* = 1.97×10^-26^), which are directly linked to aging and myocardial structure and function. Imaging-specific pathways included system-level and disease pathways, such as oxidative phosphorylation (*P* = 4.15×10^-116^), non-alcoholic fatty liver disease (*P* = 1.20×10^-86^), Alzheimer’s disease (*P* = 1.38×10^-71^), and hypertrophic cardiomyopathy (*P* = 1.52×10^-22^). Within shared pathways, protein-based aging clock of the heart had stronger enrichment in cellular-level pathways, such as estrogen signaling pathway (*P* = 4.35×10^-47^) and PI3K-Akt signaling pathway (*P* = 7.26×10^-16^). Meanwhile, the imaging-based aging clock of the heart showed stronger enrichment in neurodegenerative and cardiovascular disease pathways, such as diabetic cardiomyopathy (*P* = 1.81×10^-92^), Parkinson disease (*P* = 4.77×10^-85^), and amyotrophic lateral sclerosis (*P* = 1.36×10^-73^). Aging clocks for the brain measured by protein and imaging modalities showed consistent patterns. Specifically, gene modules for brain sMRI, dMRI, and rfMRI were enriched in 23, 112, and 149 pathways, respectively. In contrast, the protein-based aging clock was enriched in 168 pathways, with 113 of these overlapping with imaging-based aging clocks. Brain sMRI and dMRI aging clocks were specifically enriched in neurodegenerative disease pathways, such as Parkinson’s disease (*P* < 7.58×10^-26^) and Alzheimer’s disease (*P* < 5.54×10^-19^). Protein-based aging clocks showed greater enrichment in cellular-level pathways, such as endocytosis (*P* < 6.36×10^-28^) and regulation of actin cytoskeleton (*P* < 4.15×10^-14^). Brain rfMRI and protein-based aging clocks shared more cellular-level pathways, including PI3K-Akt signaling (*P* < 1.86×10^-10^) and ECM-receptor interaction (*P* < 7.41×10^-33^). Detailed results for other organs and pleiotropic gene modules across multiple organs are provided in the Supplementary Note.

When examining organ-wise specificity within each modality, we found that imaging-based aging clocks had a much higher number of organ-specific pathways than protein-based clocks (**Fig. 4C**). This likely reflects the strong ability of imaging traits to capture localized structural and functional changes. For example, unsaturated fatty acid synthesis and alpha-linolenic acid metabolism were uniquely enriched in the brain sMRI aging clock, likely representing the brain’s reliance on long-chain polyunsaturated fatty acids such as DHA to maintain membrane integrity, synaptic function, and neuroimmune homeostasis^60,61^. Additionally, drug metabolism and tryptophan metabolism were uniquely enriched in imaging-based liver aging clock, consistent with the liver’s central role in xenobiotic detoxification and tryptophan catabolism, mediated by enzymes such as cytochrome P450s and indoleamine dioxygenases^62,63^ (**Fig. 4D**).

We next focused on 15 KEGG pathways previously implicated in aging^64–76^ and were frequently enriched across multiple aging clocks (average frequency = 8.25 clocks) (**Fig. 4E**). We classified these pathways into five functional categories, including neurodegeneration, cancer, stress response and DNA damage, mitochondria and energy, as well as inflammation and immunity. Notably, imaging-based and protein-based liver aging clocks were associated with nearly all these 15 pathways (14 and 15, respectively). In addition, protein-based aging clocks of the brain, heart, lung, and adipose were enriched in over 11 of these pathways, while imaging-based clocks for the brain, artery, and kidney showed enrichment in more than 10. Therefore, aging clocks from different organs and modalities were often enriched in the same aging-related biological process and disease pathway. Since most of their gene modules were context-specific (**Fig. 3B**), these results suggest that aging clocks from different organs often converge and jointly capture well-established pathways by representing distinct fragments of the underlying complex aging process. This underscores the complementary roles of organ-specific and modality-specific aging clocks in capturing biological and disease mechanisms.

We provided further biological insights into three pathways (**Fig. 4F**). For example, 11 protein-based and imaging-based aging clocks were enriched in the KEGG Alzheimer’s disease pathway (*P* < 2.12 ×10^-4^), including both organ-specific gene modules and pleiotropic modules spanning multiple organs, together covering 65.7% of genes involved in this pathway. Several aging clocks captured distinct components of the Alzheimer’s disease pathway. For example, the brain dMRI aging clock was enriched through *FZD* and *WNT* family genes, which have been implicated in Alzheimer’s disease^77–80^. In contrast, the brain sMRI aging clock was linked to *DVL2* and *NAE1*^81,82^, while the enrichment of brain protein-based aging clock was related to *MAP2K1* and *RAF1*^83,84^. Specific pathway enrichments were also observed in aging clocks of other organs, including the liver, kidney, and adipose. Additionally, shared components enriched across multiple aging clocks were observed, primarily involving genes from the *NDUF*, *UQCR*, *PSM*, *COX*, and *TUB* families^85,86^. Similarly, we identified 15 and 11 aging clocks enriched in the KEGG cancer and NF-kappa B signaling pathways, encompassing 57.2% and 68.6% of the genes within these pathways, respectively. Both specific and shared pathway enrichments were observed, involving genes with well-documented relevant functions, such as *APC*, *KRAS*, *PTEN*, *RB1*, and *TP53*^87–91^. These examples illustrate how modality-specific and organ-specific aging clocks can work together to dissect broader biological pathways, with each clock reflecting unique and complementary insights into the complex and systemic nature of the aging process.

### Pan-organ aging clocks associated with physical activity and sleep

Physical activity and sleep are key modifiable lifestyle factors influencing healthy aging and age-related diseases. We examined genetic and phenotypic correlations between modality-specific and organ-specific aging clocks and various categories of physical activity measures and sleep phenotypes (**Methods**).

At a 5% FDR level for both genetic and phenotypic correlations, modality-specific aging clocks were associated with various physical activities (*P* < 4.6×10^-3^; **Figs. 5A-5C** and **Tables S6-S9**). Phenotypically, all aging clocks, regardless of whether they were imaging-based or protein-based, were associated with three categories of physical activity traits: sedentary behavior, circadian rhythm, and overall activity volume (Fig. 5A). On the other hand, genetic associations were primarily observed with imaging-based aging clocks. Notably, imaging-based aging clocks of the adipose and muscle showed a wide range of genetic associations with physical activity (**Figs. 5B-5C**). We also observed imaging-based bone aging clock showing consistent positive associations with several physical activity traits (*P* < 4.6×10^-3^), suggesting that certain types of physical activity may have unintended adverse effects on bone health. In contrast, among protein-based aging clocks, only the lung aging clock showed multiple genetic associations with physical activity traits. These included negative correlations with faster walking pace, greater time spent in moderate activity, and participation in strenuous sports, as well as positive correlations with longer walking duration and increased time spent watching television, which were consistent with findings in previous studies^92–95^.

**Fig. 5.**
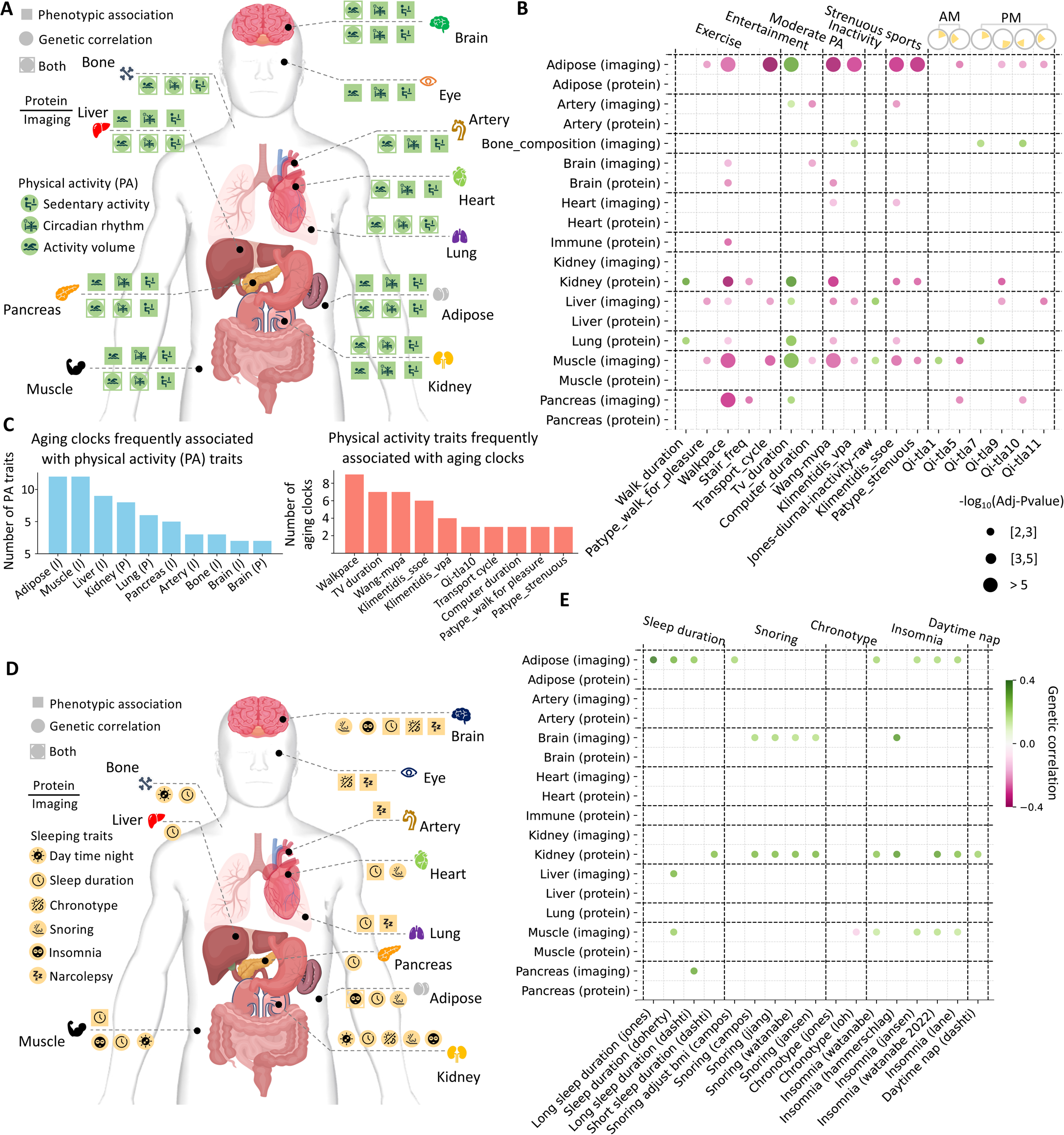
Associations between lifestyle factors and aging clocks. (**A**). Overview of genetic and phenotypic correlations between aging clocks and physical activity traits with false discovery rate (FDR)-adjusted *P*-values < 0.05. Squares indicate significant phenotypic correlations, while circles represent significant genetic correlations. Symbols above the dashed line correspond to protein-based aging clocks, and those below represent imaging-based clocks. (**B**). Selected genetic correlations between aging clocks and physical activity traits. Detailed descriptions of physical activity traits are provided in **Table S31**. (**C**). Aging clocks most frequently associated with physical activity traits (left) and physical activity traits most frequently associated with aging clocks (right), based on genetic correlation results. (**D**). Overview of genetic and phenotypic correlations between aging clocks and sleep traits with FDR-adjusted *P*-values < 0.05. Squares indicate significant phenotypic correlations, while circles represent significant genetic correlations. Symbols above the dashed line correspond to protein-based aging clocks, and those below represent imaging-based clocks. (**E**). Selected genetic correlations between aging clocks and **sleep traits. Detailed descriptions of sleep traits are provided in Table S32.**

We also identified several physical activity traits that were widely associated with aging clocks in multiple organs (**Figs. 5B-5C**). For example, longer TV duration was consistently associated with increased aging in the adipose, artery, kidney, liver, lung, muscle, and pancreas (*P* < 4.6×10^-3^), and similarly, inactivity was also positively associated with aging in the liver and muscle. In contrast, several active behaviors were consistently linked to reduced aging across organs. Particularly, a faster walking pace was associated with slower aging in the adipose, brain, immune system, kidney, liver, lung, muscle, and pancreas. Similarly, higher stair climbing frequency was protective for the pancreas and kidney aging, cycling for transport was linked to reduced aging in the liver and muscle, and moderate to strenuous physical activity showed widespread benefits across the adipose, artery, brain, heart, immune system, liver, kidney, muscle, and pancreas (*P* < 2.22×10^-3^). In addition, timing of physical activity showed distinct effects. Higher physical activity intensity between 8-10 AM (measured by the Qi-tla5 trait^96^, derived from wrist-worn accelerometer) was positively associated with reduced aging in the adipose, muscle, and pancreas. A similar effect was observed for higher physical activity intensity between 4-6 PM (Qi-tla9), but in the adipose, kidney, and liver^97–99^. In contrast, higher physical activity intensity between 12-2 AM (Qi-tla1) or 12-2 PM (Qi-tla7) was associated with increased aging in the muscle and lung, respectively.

We also observed modality-specific association patterns between aging clocks and sleep traits and disorders (*P* < 4.6×10^-3^), with genetic and phenotypic correlations exhibiting distinct patterns (**Figs. 5D-5E** and **Tables S10-S13**). Among these imaging-based aging clocks, only adipose was significantly associated with insomnia at both the genetic and phenotypic levels^100^. Genetic analyses revealed that snoring was associated with aging clocks of brain dMRI, brain rfMRI, and adipose (*P* < 2×10^-3^), while insomnia showed links to brain rfMRI, adipose, and muscle aging clocks (*P* < 2.7×10^-3^). Sleep duration was also genetically correlated with aging clocks of bone composition, pancreas, and liver (*P* < 3.4×10^-3^). On the phenotypic level, additional associations were observed between eye aging clocks and both chronotype and narcolepsy (*P* < 9.2×10^-3^). In contrast, protein-based aging clocks exhibited fewer associations, with genetic findings mostly related to the kidney. Specifically, the kidney aging clock showed significant genetic links to multiple sleep phenotypes, including snoring, daytime napping, insomnia, and sleep duration (*P* < 8×10^-4^). This link may reflect shared mechanisms reported in the literature, including disruptions in fluid balance, hormonal regulation, and autonomic function^101,102^. Phenotypic associations of protein-based aging clocks were more limited, with narcolepsy linked to artery and lung aging clocks, and sleep duration associated with muscle and lung aging clocks (*P* < 1.0×10^-2^).

### Implications of aging clocks in genetic risk for clinical endpoints

We used curated GWAS data resources on clinical endpoints from the FinnGen study^103^ to investigate genetic correlations and putative causal genetic relationships in Mendelian randomization (MR) between modality-specific and organ-specific aging clocks and diseases (**Methods**). At a 5% FDR level for genetic correlations (*P* < 1.0×10^-2^) and using a stringent Bonferroni correction for MR associations (*P* < 4.36×10^-6^), we observed modality-specific association patterns between aging clocks and diseases (**Fig. 6A** and **Tables S14-S19**).

**Fig. 6.**
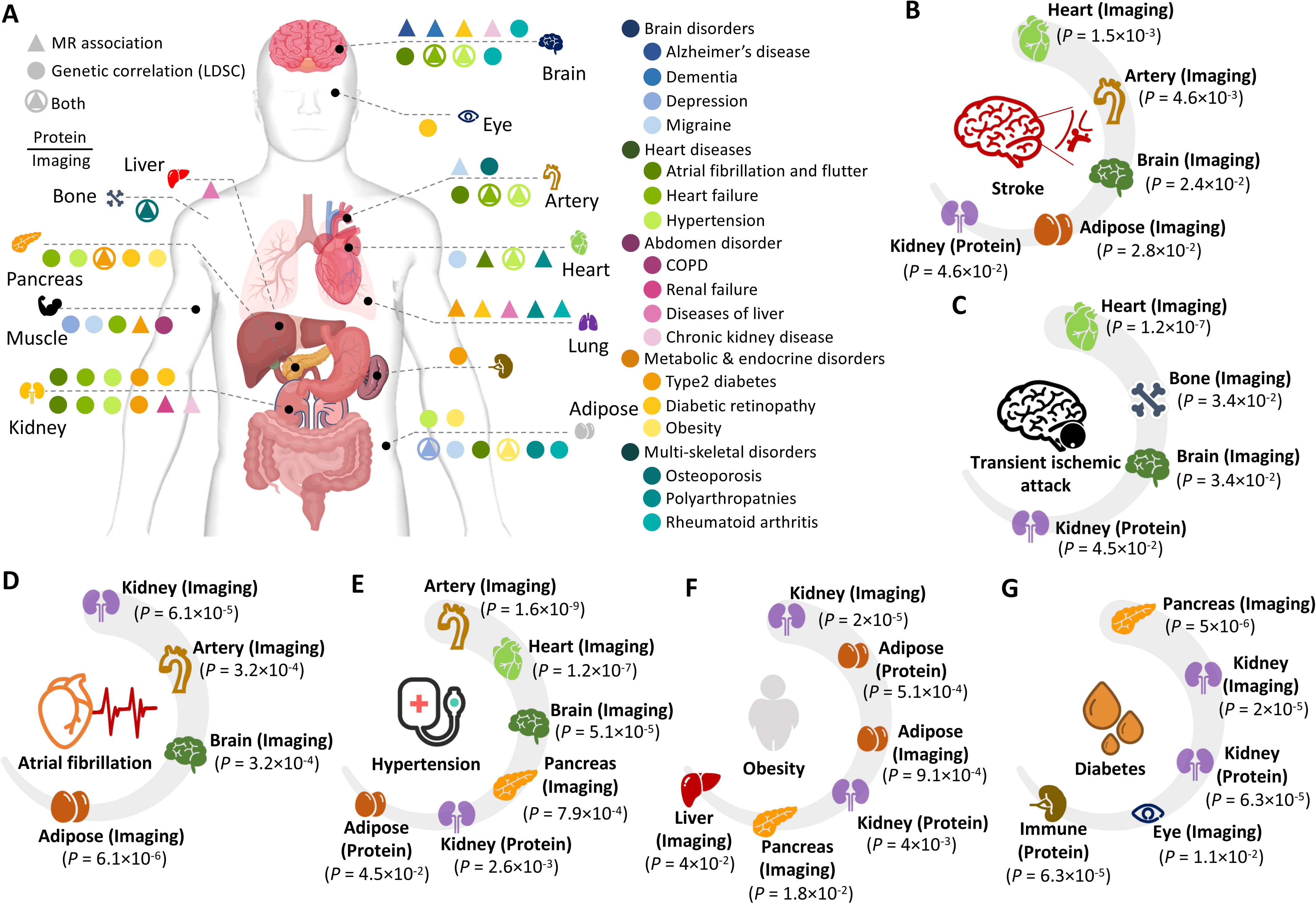
Genetic associations between clinical endpoints and aging clocks. **(A**). Overview of LDSC-based genetic correlations and Mendelian randomization (MR)-based putative causal genetic associations between aging clocks and physical activity traits after multiple testing adjustments. Triangles indicate MR associations, while circles represent genetic correlations. We annotate a MR association if either causal direction is significant. Symbols above the dashed line correspond to protein-based aging clocks, and those below represent imaging-based clocks. (**B**)-(**G**). Examples of diseases that showed genetic correlations with aging clocks across multiple organs, including brain disorders (stroke (**B**) and transient ischemic attack (**C**)), cardiovascular diseases (atrial fibrillation (**D**) and hypertension (**E**)), and systemic conditions (obesity (**F**) and type 2 diabetes (**G**)).

Several imaging-based aging clocks demonstrated consistent links to multiple diseases across both genetic correlations and MR relationships. These included type 2 diabetes with the aging clock of the pancreas, osteoporosis with the aging clock of the bone, heart failure with aging clocks of both the artery and brain, hypertension with aging clocks of the artery, heart, and brain, and obesity with aging clock of adipose (*P* < 3.40 × 10^-3^). Specifically, the kidney clock showed genetic correlations with several cardiovascular diseases and had MR associations with both renal failure and chronic kidney disease. The adipose aging clock was genetically correlated with brain disorders, heart diseases, and musculoskeletal conditions. The pancreas aging clock was genetically linked to both metabolic disorders such as type 2 diabetes and cardiovascular diseases including heart failure and hypertension. Protein-based aging clocks of the kidney, brain, and lung also showed notable genetic associations. Like imaging-based kidney aging clock, the protein-based kidney aging clock had genetic correlations with cardiovascular conditions such as hypertension and atrial fibrillation, as well as metabolic disorders, such as type 2 diabetes. The protein-based lung aging clock had MR associations with multiple diseases, including type 2 diabetes, liver diseases, and musculoskeletal disorders. For the brain, the protein-based aging clock showed MR associations with Alzheimer’s disease, dementia, and chronic kidney disease.

Consistent with findings from our pathway analysis (**Figs. 4D-4E**), many diseases exhibited genetic associations with aging clocks from both imaging and protein modalities across multiple organs. For example, stroke was genetically associated with imaging-based aging clocks of the heart, artery, brain, and adipose, as well as the protein-based kidney aging clock (**Fig. 6B**). Similarly, transient ischemic attack showed associations with imaging-based aging clocks of the heart, bone composition, and brain, and with the protein-based kidney aging clock (**Fig. 6C**). The connections between these brain disorders and the heart aging clock may be due to cardioembolic phenomena, increased large artery stiffness and/or abnormal regulation of cerebral blood flow ^104^, as well as contributions from small vessel disease and thrombo-embolic events^105,106^. Aging in the adipose may promote chronic inflammation and metabolic dysfunction, and kidney aging affects blood pressure regulation and vascular health, all of which elevate risk of brain-related diseases^107–110^. These aging clocks may capture distinct biological aging components that underlie the genetic risk of brain diseases.

Cardiovascular diseases also showed associations with aging in multiple organs from different modalities. Atrial fibrillation was linked to imaging-based aging clocks of the kidney, artery, brain, and adipose (**Fig. 6D**), while hypertension was associated with imaging-based clocks of the artery, heart, brain, and pancreas, as well as protein-based clocks of the kidney and adipose (**Fig. 6E**). These associations may reflect how aging in peripheral organs contributes to cardiovascular diseases through mechanisms such as impaired vascular integrity, large artery stiffening (which promotes excessive pulsatile load to the left ventricle, contributing to ventricular remodeling, heart failure and atrial arrhythmias), disrupted fluid and blood pressure regulation, metabolic dysfunction, and systemic inflammation^111–114^. In addition to brain and heart-related disorders, several systemic conditions showed broad associations with multiple aging clocks. For example, obesity was associated with imaging-based aging clocks of the kidney, adipose, pancreas, and liver, as well as protein-based clocks of the adipose and kidney (**Fig. 6F**). These genetic correlations may reflect disruptions in fat storage, insulin regulation, metabolic homeostasis, and fluid balance, core systems underlying energy metabolism^115–118^. Moreover, type 2 diabetes was linked to imaging-based clocks of the pancreas, kidney, and eye, and to protein-based clocks of the kidney and immune system, reflecting the central roles these organs play in glucose regulation, insulin production, vascular complications, and immune dysfunction commonly observed in diabetes^119–121^ (**Fig. 6G**).

## DISCUSSION

This study presents a comprehensive resource for dissecting the molecular and structural dimensions of biological aging across multiple organs, using protein-based and imaging-based aging clocks constructed from the large-scale UKB study. Our analyses revealed distinct yet complementary mechanistic insights from the derived modality-specific and organ-specific aging clocks, demonstrating how jointly considering these aging signatures enhances our understanding of context-specific aging trajectories and their connections to health outcomes and lifestyle factors.

One of our novel contributions is the characterization of distinct phenotypic and genetic signatures between imaging-based and protein-based aging clocks, along with the observed contrast in the alignment between phenotypic and genetic correlations across aging clocks within different data modalities. Imaging-based aging clocks exhibited strong coherence between genetic and phenotypic correlations, suggesting that heritable factors substantially shape structural and physiological aging trajectories. In contrast, protein-based aging clocks, while phenotypically correlated, showed far fewer and weaker genetic correlations. This divergence may reflect the nature of proteomic aging measures, which are sensitive to environmental and lifestyle influences and may capture early molecular perturbations, such as inflammation or metabolic dysregulation, that precede heritable long-term structural decline^41,122^. Moreover, protein aging clocks may reflect dynamic pathways subject to short-term changes, including not only pathways that promote accelerated aging, but also counterregulatory pathways, whereas imaging aging clocks represent the net effect long-term effect of pathways that promote structural organ aging. Consequently, a greater proportion of individuals were flagged as organ agers using protein-based aging clocks^42,123^. Across both modalities, approximately 2% of individuals were classified as extreme agers in each organ, while about 5% showed evidence of multi-organ aging. Surprisingly, only 0.3% exhibited aging in the same organ across both imaging-based and protein-based aging clocks, but this overlap rose to 2.6% among multi-organ agers, indicating more systemic and advanced biological aging. Notably, aging was more frequently observed in metabolically active and detoxification-related organs, such as the liver, kidney, pancreas, spleen, and adipose. These organs are especially vulnerable to cumulative physiological stress, which may explain their early and consistent involvement in individuals with widespread aging.

At the individual-level, we observed that individuals sharing accelerated aging in the same organ often exhibited divergent aging patterns in other organs. Such variations may contribute to the low genetic and phenotypic correlations across individuals at the cohort-level. To better understand this variability, we examined the genetic underpinnings of different aging clocks and found that, despite both being heritable, protein-based and imaging-based clocks are shaped by largely distinct genetic architectures. In network biology analysis, we identified largely different gene modules across aging clocks, with minimal shared components even for imaging-based and protein-based aging clocks measuring the same organ. While most gene modules exhibited context specificity, we identified a small number of pleiotropic gene modules shared between aging clocks from different organs and modalities. For example, shared modules between imaging-based aging clocks of the brain and artery highlighted connections between disparate organs both within and across modalities. These findings provide a multi-organ perspective on systemic aging and may help uncover core biological pathways and therapeutic targets that contribute to aging-related decline across the brain and body.

We further conducted a systematic pathway enrichment analysis for all gene modules. We found that protein-based clocks were more specifically enriched in cellular related pathways, whereas imaging-based aging clocks showed stronger enrichment in disease and system level pathways, such as oxidative phosphorylation. More importantly, these aging clocks often converged and jointly captured established aging and disease pathways. Notably, the KEGG Alzheimer’s disease pathway was enriched across gene modules from multiple aging clocks, including brain imaging-based aging clocks (sMRI, dMRI, and rfMRI), protein-based aging clocks for the brain, liver, and kidney, as well as gene modules shared across multiple organs. Each aging clock contributed partially different sets of genes, together covering over 65.7% of the whole KEGG Alzheimer’s disease pathway. These findings highlight how modality-specific and organ-specific aging clocks capture complementary parts of complex disease biology, which could help enable more precise dissection of disease mechanisms and support the development of targeted, multi-organ interventions and therapeutic strategies for age-related disorders like Alzheimer’s disease.

In addition to biological mechanisms, we explored how modality-specific aging clocks relate to physical activity and sleep, well-established modifiable lifestyle factors. Imaging-based aging clocks, particularly those for the brain, adipose, and muscle, showed genetic associations with sleep traits such as insomnia, snoring, and sleep duration. At the phenotypic level, brain imaging clocks were also associated with chronotype and narcolepsy, suggesting that structural aging in the brain may relate to sleep regulation. In contrast, protein-based aging clocks showed fewer associations with sleep overall; however, the kidney clock stood out by displaying consistent genetic and phenotypic links to multiple sleep-related phenotypes, such as snoring, daytime napping, insomnia, and sleep duration This may reflect the kidney’s central role in maintaining systemic physiological balance, which can impact sleep-related processes. However, insomnia and sleep disordered breathing can also impact blood pressure, sympathetic activity, and metabolic regulation, which can in turn have downstream effects on kidney aging. The cause effect relationships of the observed associations require further study.

Physical activity showed widespread and modality-specific associations with biological aging. Phenotypically, nearly all aging clocks, across both imaging and protein modalities, were associated with sedentary behavior, circadian rhythm, and overall activity volume. Genetically, these associations were most prominent for imaging-based clocks, particularly those of the adipose tissue and muscle. In contrast, protein-based clocks showed fewer genetic associations, with the lung aging clock being a notable exception, which had several connections with physical activity traits, such as moderate to strenuous activates. Among physical activity traits, longer TV duration consistently emerged as a strong factor of accelerated aging in multiple organs, especially the liver, kidney, pancreas, muscle, and adipose, based primarily on imaging-derived aging clocks. Timing of physical activity also played a critical role: early morning (8-10 AM) and later afternoon (4-6 PM) activities were linked to reduced aging in muscle and some abdominal organs, whereas late-night (12-2 AM) and midday (12-2 PM) activities were associated with increased aging in the lung and muscle. Furthermore, faster walking pace, higher stair climbing frequency, higher cycle frequency and moderate-to-strenuous exercise were consistently associated with reduced aging across multiple organs. These findings suggest that the intensity, duration, and timing of physical activity all influence biological aging in an organ-specific manner. Encouraging healthier physical activity pattern throughout the day may help slow biological aging across multiple organs.

To assess the clinical implications of biological aging, we evaluated genetic correlations and MR relationships between aging clocks and disease outcomes using data from the FinnGen study^103^. Imaging-based aging clocks showed stronger and more consistent associations with multiple major diseases, such as type 2 diabetes (pancreas aging clock), heart failure and hypertension (artery and brain), and Alzheimer’s disease (adipose). These findings highlight key inter-organ connections, including the brain-heart axis^124,125^ and metabolic-neurodegenerative axis^126,127^. Protein-based clocks revealed fewer associations overall but also identified biologically meaningful links. For example, the kidney protein-based aging clock was associated not only with cardiovascular conditions, like its imaging-based counterpart, but also with chronic kidney disease and renal failure. Similarly, the brain protein-based aging clock showed strong MR associations with Alzheimer’s disease and dementia. These findings suggest that protein-based clocks may detect early molecular alterations that precede structural decline, offering complementary value to imaging-based measures in identifying silent or subclinical stages of disease. Notably, several common and clinically important diseases, including stroke, atrial fibrillation, hypertension, obesity, and type 2 diabetes, showed genetic associations with aging clocks across multiple organs and both data modalities. These findings suggest that aging clocks across different organs may reflect disease risk through distinct yet complementary mechanisms, underscoring the importance of viewing aging as a coordinated multi-organ process.

Together, our findings illustrate the strengths of molecular and structural aging clocks in revealing organ-specific and systemic patterns of aging, their underlying biological mechanisms, and clinical implications. To our knowledge, this study represents the largest cohort to date integrating multi-organ imaging and proteomic data. By integrating these multi-modal aging measures, we provide a foundational resource for precision aging research and provides potential avenues for early intervention, risk stratification, and personalized medicine.

## METHODS

Methods are available in the ***Methods*** section. Note: One supplementary PDF file and one supplementary Excel file are available.

## ACKNOWLEDGEMENTS

Research reported in this publication was supported by National Institute of Mental Health under Award Number R01MH136055 and National Institute on Aging under Award Numbers RF1AG082938 and R01AG085581. The content is solely the responsibility of the authors and does not necessarily represent the official views of the National Institutes of Health. The study has also been partially supported by funding from the Purdue University Statistics Department, Department of Statistics and Data Science at the University of Pennsylvania, Wharton Dean’s Research Fund, Analytics at Wharton, Wharton AI & Analytics Initiative, Perelman School of Medicine CCEB Innovation Center Grant, and the University Research Foundation at the University of Pennsylvania. The individual-level data from the UK Biobank study were obtained under application 76139 subject to a data transfer agreement. This research has been conducted using summary-level data from the FinnGen research project and other studies that made their summary-level data publicly available. We would like to thank the individuals who represented themselves in the UK Biobank, FinnGen, and other related studies for their participation and the research teams for their efforts in collecting, processing, and disseminating these datasets. We would like to thank the research computing and IT groups at the Wharton School of the University of Pennsylvania and the Rosen Center for Advanced Computing at the Purdue University for providing computational resources and support that have contributed to these research results.

## AUTHOR CONTRIBUTIONS

J.S. and B.Z. designed the study. J.S. performed the data analysis and generated the results. Y.G., Z.F., Y.Y., Y.L., X.Y., C.B., I.H., B.S., and R.Z. contributed to data preprocessing, analysis, and interpretation of results. J.C., P.P., L.W., J.M.O., R.G., J.L., W.W., D.J.R., and A.R. provided input on the research question, study design, and interpretation of findings. J.S. and B.Z. wrote the manuscript with feedback from all authors.

## METHODS

### UK Biobank imaging data

The multi-organ imaging data used in this study were from the UK Biobank (UKB), a large-scale biomedical database that recruited around 500,000 participants aged 40 to 69 between 2006 and 2010 (https://www.ukbiobank.ac.uk/). The majority of these imaging data, spanning multiple organs, are derived from the ongoing UKB imaging initiative (https://www.ukbiobank.ac.uk/explore-your-participation/contribute-further/imaging-study). This initiative aims to collect brain, cardiac, and abdominal magnetic resonance imaging (MRI), along with dual-energy X-ray absorptiometry (DXA) scans, from up to 100,000 individuals. The UKB study has received ethical approval from the North West Multicentre Research Ethics Committee (reference: 11/NW/0382).

Brain MRI provided comprehensive insights into both structural and functional aspects of the brain^128^. We used image-derived phenotypes (IDPs) generated by the UKB brain imaging pipeline^129^ (https://biobank.ndph.ox.ac.uk/ukb/label.cgi?id=508), including 258^130^ regional brain volume metrics from structural MRI (sMRI), 432 microstructural white matter properties from diffusion MRI (dMRI), and 1,777 functional connectivity/activity traits from resting-state functional MRI (rfMRI) (**Tables S20-S22**). In addition, we used 82 traits^131–133^ generated from cardiovascular magnetic resonance imaging to assess cardiac and aortic structure and function (https://biobank.ndph.ox.ac.uk/ukb/label.cgi?id=157). These traits encompass measurements of the left and right ventricles, left and right atria, and the ascending and descending aorta (**Table S23**). Abdominal IDPs included 41 MRI-derived traits encompassing body fat, muscle composition, and organ-specific features (https://biobank.ndph.ox.ac.uk/ukb/label.cgi?id=105). There were 13 traits quantifying abdominal fat distribution and visceral/subcutaneous fat volumes, and 7 traits assessing muscle characteristics, including lean and fat-free muscle volume in the thighs and muscle fat infiltration as a marker of muscle quality. Organ-level measures included 6 kidney traits (volume and parenchyma), 10 liver traits (fat and iron content, with corrected T1 reflecting inflammation and fibrosis), 3 pancreas traits (volume, fat, and iron), as well as single-volume traits for the lung and spleen (**Table S24**).

The DXA-derived traits provided 71 body composition measurements, capturing the distribution of fat, lean mass, and tissue composition across multiple body regions (https://biobank.ndph.ox.ac.uk/ukb/label.cgi?id=124). These traits included fat-free mass and tissue mass in the android and gynoid regions, as well as in specific limbs such as the left/right arm and leg). Additional measures included total body fat mass, lean mass, and bone mass, offering a comprehensive overview of body composition (**Table S25**). In addition, we included 119 bone composition traits assessed via DXA scans, quantifying bone mineral content, bone mineral density, and bone area across key skeletal sites such as the lumbar spine, femoral neck, and total body (https://biobank.ndph.ox.ac.uk/ukb/label.cgi?id=125). There were also hip shape mode scores and osteophyte gradings, which reflected bone geometry and joint degeneration, respectively (**Table S26**).

We also used 156 retinal imaging features^134^, including 46 derived from optical coherence tomography (OCT) and 110 extracted from fundus photographs. The OCT-based metrics captured detailed structural information such as retinal layer thicknesses, vertical cup-to-disc ratio, and optic disc diameter. For the fundus images, 11 pre-trained deep learning models originally trained on ImageNet were applied, and from the final layer of each model, the top 10 principal components (PCs) were extracted, yielding a total of 110 distinct fundus image features (**Tables S27-S28**). For each IDP, outliers were excluded, defined as values exceeding five times the median absolute deviation from the median^135^.

### UK Biobank pharma proteomics project

The UK Biobank pharma proteomics project (UKB-PPP) analyzed plasma proteomic profiles from 54,219 participants using the Olink Explore 3072 library. This high-throughput assay measured 2,941 protein analytes corresponding to 2,923 unique proteins across eight targeted panels related to inflammation, cardiometabolic function, neurology, and oncology (**Table S29**). Protein abundance was reported as normalized expression values and processed via Olink’s MyData Cloud platform. Additional details regarding sample handling, processing, and quality control are available in a previous study^136^.

We annotated plasma proteins with organ-specific labels using a method adapted from Oh et al.^14^. In brief, gene expression data were sourced from GTEx v8^137^, with expression levels normalized across tissues. A gene, and by extension, its corresponding protein, was labeled as organ-specific if its expression in a single organ was at least fourfold higher than in any other organ, following the criteria established by the Human Protein Atlas. To maintain consistency across similar tissues, sub-tissues belonging to the same organ were grouped, and the highest expression value among them was used to represent that organ. As in Oh et al.^14^, the immune organ category was defined by aggregating gene expression from blood and spleen tissues. Applying this method, we assigned organ-specific labels to 557 proteins based on their gene expression signatures (**Table S30**).

### Lifestyle factor traits

The individual-level physical activity traits were collected from the touchscreen questionnaire and accelerometer-derived data in the UKB study (**Supplementary Note**). Genome-wide association studies (GWAS) summary statistics of physical activity traits were obtained from multiple previous literature involving UKB^138–143^. We categorized physical activity traits into three groups: activity volume, sedentary behaviors, and circadian rhythm. For the phenotypic correlation analysis, we included 79 physical activity traits that we had individual-level data, consisting of 42 touchscreen-assessed traits and 37 accelerometer-measured traits. These were a subset of the 83 physical activity traits for which we had GWAS summary statistics, and which were therefore also used in the genetic correlation analysis. Detailed descriptions and sources of these physical activity traits can be found in **Table S31.** Similarly, we used 34 publicly available sleep GWAS datasets, covering seven distinct sleep traits (**Table S32**). For the phenotypic correlation analysis, we examined 10 sleep traits with available individual-level data, including seven traits based on touchscreen questionnaire responses and three traits derived from accelerometer data. Additional details about these data are provided in the **Supplementary Note**.

### FinnGen clinical endpoints

We analyzed 372 clinical endpoints from the FinnGen project (R9 release; https://www.finngen.fi/en/access_results), including 304 conditions with more than 10,000 cases and an additional 68 with at least 5,000 cases (**Fig. S3**). To ensure adequate statistical power for Mendelian randomization (MR), we performed a two-sample MR power analysis (**Supplementary Note** and **Fig. S4**), using 10,000 cases as a general threshold for inclusion^144^. While many endpoints met this criterion, a few with smaller sample sizes, such as Alzheimer’s disease, was retained due to their biological relevance to aging. There are no universally accepted standards for the minimum number of cases required in two-sample MR studies. For example, common diseases like hypertension often involve over 10,000 cases^145^, whereas uncommon diseases can have less than 1,000 cases. The 372 clinical endpoints span a wide array of disease categories, including psychiatric, circulatory, respiratory, digestive, genitourinary, endocrine, autoimmune, and musculoskeletal disorders. Endpoint definitions are accessible at https://risteys.finregistry.fi/. Notably, the FinnGen data used in this analysis were derived from cohorts independent of the UKB study, avoiding any potential sample overlap. Full details of the included endpoints are provided in **Table S33**.

### Deriving modality-specific and organ-specific aging clocks

Following the procedure described in Oh et al.^14^, we used organ-specific imaging or protein data to predict the chronological age using least absolute shrinkage and selection operator (LASSO) regression. The predicted chronological age was then compared to the actual age by fitting a linear regression between the two. Individual age gaps were calculated as the difference between each person’s predicted age and the linear regression estimate of the population mean. This gap between predicted biological age and actual chronological age provides a personalized estimate of biological aging. We used unrelated individuals of white ancestry^146^ for this analysis. We also assessed feature importance from the LASSO models. To evaluate the stability of each feature, we performed 100 bootstrap iterations using the original sample size. For each feature, we calculated the average effect size and the proportion of times it was retained (that is, not shrunk to zero) across all iterations (**Figs. S5-S6)**. In total, we constructed 16 imaging-based and 10 protein-based aging clocks, including paired clocks for 10 major organs (**Table S34**). We then excluded two imaging-based clocks (lung and immune) due to their low prediction accuracy and small number of stable predictors (model prediction *R*^2^ < 0.2) and used the remaining 24 aging clocks (including paired clocks for 8 organs) for all downstream analyses.

### Discovery and replication GWAS of aging clocks

We performed GWAS separately for aging clocks using linear mixed models via fastGWA^147^. Aging clocks were all standardized by subtracting the mean and dividing by the standard deviation. Imputed genotype data from the UKB study were utilized, following standard quality control procedures consistent with our prior research^125^. Specifically, we excluded individuals with genotype missingness > 10%, removed variants with missingness >10%, filtered out variants with minor allele frequency < 1%, discarded those failing Hardy-Weinberg equilibrium at *P* < 1×10^-7^, and excluded variants with imputation INFO scores below 0.8. The GWAS models included covariate adjustments for standing height, BMI, age (at data collection), sex, assessment center, age squared, age and sex interaction, and the top 40 genetic PCs^146^. The aging clock based on brain rfMRI imaging traits was additionally adjusted for the head motion variables^125^.

We used unrelated individuals of white ancestry for the GWAS analysis and randomly split the data into independent discovery and replication cohorts of equal size (average discovery sample size *n* = 21,019 for imaging and 20,800 for proteins). For the discovery samples, we identified variants significantly associated with aging clocks (*P* < 5×10^-8^/24) and clumped them based on linkage disequilibrium (LD) using the publicly available tool python_convert (https://github.com/precimed/python_convert), which is an API version of the FUMA^43^ procedure and integrates PLINK and an LD reference panel from the European population of the 1000 Genomes Project Phase 3. Independent significant variants were retained if they had pairwise LD *r*^2^ < 0.1. We then assessed the replicability of these independent variants by examining their effect sizes and significance levels in the independent replication cohort. We found that their effect sizes all had consistent directions in the two cohorts (**Figs. S1-S2**). Replicated GWAS signals were then categorized into four levels of confidence: (i) Extremely high confidence, if they were genome-wide significant in the replication sample after Bonferroni correction for the number of aging clocks (*P* < 5×10^-8^/24); (ii) High confidence, if they passed the standard genome-wide significance threshold (*P* < 5×10^-8^); (iii) Moderate confidence, if they reached a nominal significance level adjusted for the number of independent variants in the discovery sample (*P* < 0.05/1336); and (iv) Low confidence, if they did not meet any of the above thresholds. Variants with extremely high confidence were mapped to chromosomal bands and visualized across the genome (**Fig. 3A**).

We used GCTA (v1.93.2) to estimate the heritability of each trait under a linear mixed model framework^148^. We used data from all unrelated individuals of white ancestry, and the analyses were adjusted for the same set of covariates as in the GWAS analysis.

### Identifying seed genes of aging clocks

Based on the high replication rate observed in GWAS signals of the aging clocks, we conducted a GWAS on the combined discovery and replication samples to enhance statistical power for subsequent network analysis. To prepare seed genes for network expansion, we performed gene-based analysis of our aging clocks on the combined samples using MAGMA (v1.10) under the default settings^149^. Only genetic variants located within the gene regions were included in the analysis (with gene window size set to zero) and the marginal effects of SNPs were averaged across the gene. Associated genes that exceeded the Bonferroni significance threshold (*P* < 0.05/number of genes) were identified and used in downstream network analysis.

### Active gene modules of aging clocks

To identify functionally coherent gene clusters beyond conventional GWAS loci, we used the network expansion strategy proposed by Barrio-Hernandezet al.^150^. Seed genes for each aging clock were integrated with protein-protein interactions used in Barrio-Hernandezet al.^150^ (such as STRING^151^), retaining only high-confidence interactions after multiple testing adjustment with the Benjamini-Hochberg approach. Gene modules were derived using the network propagation algorithm from the original study^150^, implemented with the publicly available code and default hyperparameters. To further investigate the patterns of overlapping gene modules identified across different aging clocks, we constructed a gene module interaction network, where each node represents a distinct gene module, and an edge indicates a significant overlap (Jaccard index > 0.7) in gene content. Using this network, we identified fully connected subgraphs (cliques), where every pair of modules was mutually overlapping. Each such clique, representing a densely interconnected set of gene modules across clocks, was termed a pleiotropic gene module.

### Pathway enrichment analysis

To investigate the biological pathways enriched in our gene modules, we conducted gene set enrichment analysis using GSEApy^152^ (v1.1.8), a streamlined tool that interfaces with the Enrichr API. We used the genes from each identified gene module as input and selected KEGG Human databases to identify significantly enriched pathways. Enrichment was assessed using Fisher’s exact test, and pathways with a Benjamini-Hochberg adjusted *P*-value < 0.05 were considered statistically significant.

### Phenotypic correlation analysis

We conducted phenotypic association analyses between 24 aging clocks and 10 sleep traits and 79 physical activity traits using available individual-level data. In total, 2,136 association pairs (89 traits × 24 clocks) were evaluated in unrelated individuals of white ancestry using pairwise univariate linear regression. All models were adjusted for commonly used covariates in the literature. For touchscreen-assessed traits, adjustments included age (at assessment), sex, assessment center, body mass index (BMI), and Townsend Deprivation Index (TDI). For accelerometer-measured traits, we adjusted for age (at device wear and measurement), sex, assessment center, season of device wear, BMI, and TDI. For each association, we reported the *P*-value, difference in means, and standard error from a two-sided *t*-test, and applied Benjamini-Hochberg correction to adjust for multiple testing across all comparisons.

### Genetic correlation through LDSC

We applied cross-trait LD score regression (version 1.0.1) (https://github.com/bulik/ldsc) to estimate pairwise genetic correlations among aging clocks, as well as between aging clocks and various lifestyle factor traits and FinnGen clinical endpoints mentioned in previous sections. The analysis used the default European LD score reference panel provided by the LDSC software, based on data from the 1000 Genomes Project.

### Mendelian randomization analysis

We investigated the putative genetic causal relationships between 24 aging clocks and 372 FinnGen clinical endpoints. Prior to conducting Mendelian randomization (MR) analyses, we performed standard quality control and preprocessing steps. Genetic instruments were selected from the exposure GWAS results using a genome-wide significance threshold of 5×10^-8^. To ensure independence among instruments, we applied LD-based clumping with a 10,000 kb window and an LD *r*^2^ threshold of 0.01, using European-ancestry reference data from the 1000 Genomes Project. Harmonization of exposure and outcome datasets was conducted using the TwoSampleMR package (https://mrcieu.github.io/TwoSampleMR/), aligning effect alleles and removing ambiguous variants to ensure consistency. Bidirectional MR analyses were then performed for each aging clock and FinnGen clinical endpoint. In the forward direction, aging clocks served as exposures and FinnGen clinical endpoints as outcomes; in the reverse direction, the roles were switched. For each exposure phenotype, LD clumping was applied to identify independent instruments, and corresponding variants were extracted from both exposure and outcome datasets to enable valid inference through MR. We assessed the performance of 8 MR methods^153^, which included Inverse variance weighted (multiplicative random effect), Inverse variance weighted (fixed effect), MR-Egger, Simple Median, Weighted Median, Weighted Mode, DIVW, GRAPPLE, and MR-RAPS^154–158^, where MR Egger was used as the pleiotropy test. Additional details on how results were summarized across different MR methods, along with descriptions of quality control procedures and sensitivity analyses, are provided in the **Supplementary Note**.

## Code availability

We made use of publicly available software and tools. The final version of our analysis code will be made freely available at Zenodo upon publication.

## Data availability

Individual-level data from the UK Biobank are available at https://www.ukbiobank.ac.uk/. Summary statistics for lifestyle factors and FinnGen clinical endpoints used in this study are publicly accessible, with detailed sources provided in the **Supplementary Tables** and **Supplementary Note**.

