## Supplementary material for "Contextualizing molecular and structural aging across human organs": supp_information

**This PDF file includes:**

- Supplementary Text
- Supplementary Figures S1-S6

**Other Supplementary Materials for this manuscript include the following:**

- Tables S1 to S34(.xlsx) (available in an Excel file)

### Supplementary Text

#### Additional results on enriched biological and disease pathways

The imaging-based artery aging clock had six gene modules enriched in 118 KEGG disease and biological pathways, while the protein-based artery aging clock had nine gene modules enriched in 111 pathways. These two aging clocks shared enrichment in 77 out of 152 pathways, such as ribosome ($P < 5.20 \times 10^{-114}$), coronavirus disease ($P < 1.31 \times 10^{-97}$), and RNA transport ($P < 7.56 \times 10^{-20}$). They also overlapped in pathways related to innate immunity and inflammation, which are closely linked to aging, such as the NF-kappa B signaling pathway ($P < 1.13 \times 10^{-12}$).  Notably, the protein-based aging clock of artery was specifically enriched in neurodegeneration pathways, such as Alzheimer's diseases ($P = 1.66 \times 10^{-44}$) and Parkinson disease ($P = 2.38 \times 10^{-28}$), as well as cancer pathways, such as breast cancer ($P = 5.67 \times 10^{-18}$) and gastric cancer ($P = 7.09 \times 10^{-18}$). The imaging-based aging clock of artery was mainly enriched in pathways closely related to inflammation.

The imaging-based adipose aging clock had seven gene modules enriched in 181 GOBP and KEGG disease pathways, while the protein-based adipose aging clock had six gene modules enriched in 72 pathways. The two aging clocks shared 50 out of 203 pathways. Examples of shared pathways included RNA degradation ($P < 1.02\times10^{-19}$), longevity regulating pathway ($P < 1.02\times10^{-5}$), ribosome biogenesis in eukaryotes ($P < 1.15\times10^{-44}$), and inflammatory mediator regulation ($P = 6.57\times10^{-8}$). The protein-based aging clock of adipose specifically enriched in pathways involving neuroactive ligand-receptor interaction ($P = 2.44\times10^{-66}$), cytokine-cytokine receptor interaction ($P = 1.77\times10^{-27}$), regulation of lipolysis in adipocytes ($P = 3.09\times10^{-12}$), which capture more cellular-level interpretations. The imaging-based aging clock was specifically enriched in system-level and disease pathways, such as oxidative phosphorylation ($P = 5.66\times10^{-152}$), Parkinson disease ($P = 1.89\times10^{-117}$), Alzheimer's disease ($P = 4.66\times10^{-103}$), and pathways of neurodegeneration ($P = 3.21\times10^{-94}$).

The imaging-based muscle aging clock had nine gene modules enriched in 111 pathways, while the protein-based muscle aging clock had five gene modules enriched in 50 pathways. The two aging clocks shared 23 out of 138 pathways. Examples of shared pathways included PI3K-Akt signaling pathway ($P < 2.10×10^{-5}$) and apelin signaling pathway ($P < 8.55×10^{-5}$). The protein-based aging clock of muscle was specifically enriched in pathways on cellular-level biological process, such as ribosome biogenesis in eukaryotes ($P = 9.37×10^{-50}$) and RNA degradation ($P = 1.62×10^{-10}$). The imaging-based aging clock of muscle was specifically enriched in pathways involving synapse, including GABAergic synapse ($P = 1.40×10^{-41}$), cholinergic synapse ($P = 1.56×10^{-39}$), serotonergic synapse ($P = 1.56×10^{-39}$), glutamatergic synapse ($P = 1.85×10^{-39}$), dopaminergic synapse ($P = 3.17×10^{-38}$) and some other system-level pathways, such as circadian rhythm ($P = 1.97×10^{-21}$).

The imaging-based pancreas aging clock had two gene modules enriched in seven pathways, while the protein-based pancreas aging clock had nine gene modules enriched in 85 pathways. The two aging clocks shared four out of 85 pathways. The shared pathways included pancreatic secretion ($P < 8.15×10^{-14}$), protein digestion and absorption ($P < 3.45×10^{-12}$), fat digestion and absorption ($P < 3.11×10^{-3}$), and renin-angiotensin system ($P < 1.60×10^{-2}$). The protein-based aging clock of pancreas specifically was enriched in pathways that are more on cellular-level biological process, such as ribosome biogenesis in eukaryotes ($P = 1.38×10^{-43}$) and lysosome ($P = 5.40×10^{-17}$). The imaging-based aging clock of pancreas was specifically enriched in metabolic pathways, including glycerolipid metabolism ($P = 8.13×10^{-4}$) and steroid biosynthesis ($P = 1.39×10^{-2}$).

The imaging-based liver aging clock had eight gene modules enriched in 180 pathways, while the protein-based pancreas aging clock had nine gene modules enriched in 185 pathways. The two aging clocks shared 113 out of 252 pathways. Among the shared pathways, protein-based liver aging clock had stronger enrichment in cellular level pathways, such as serotonergic synapse ($P = 2.93×10^{-32}$) and PI3K-Akt signaling pathway ($P = 4.26×10^{-31}$), while imaging-based liver aging clock had stronger enrichment in disease pathways, such as coronavirus disease ($P = 1.58×10^{-99}$), Alzheimer's disease ($P = 4.30×10^{-29}$), and Parkinson disease ($P = 7.71×10^{-23}$). The protein-based aging clock of liver was specifically enriched in pathways that were more on cellular-level biological process, such as neuroactive ligand-receptor interaction ($P$ = 1.38×10$^{-43}$) and chemokine signaling pathway ($P$ = 5.40×10$^{-17}$). The imaging-based aging clock of liver was specifically highly enriched in metabolic pathways, including retinol metabolism ($P$ = 4.39×10$^{-52}$), beta-Alanine metabolism ($P$ = 3.55×10$^{-40}$), histidine metabolism ($P$ = 9.35×10$^{-30}$), arginine and proline metabolism ($P$ = 3.08×10$^{-21}$), tyrosine metabolism ($P$ = 3.57×10$^{-21}$), tryptophan metabolism ($P$ = 3.74×10$^{-18}$), and pyruvate metabolism ($P$ = 1.50×10$^{-17}$).

The imaging-based kidney aging clock had six gene modules enriched in 21 pathways, while the protein-based pancreas aging clock had seven gene modules enriched in 190 pathways. The two aging clocks shared 14 out of 197 pathways. The shared pathways included cellular-level biological pathways, such as sphingolipid signaling pathway ($P$ < 1.96×10$^{-20}$) and Fc gamma R-mediated phagocytosis ($P$ < 2.43×10$^{-5}$). The protein-based aging clock of kidney was specifically enriched in several signaling pathways such as Ras signaling pathway ($P$ = 2.36×10$^{-45}$) and PI3K-Akt signaling pathway ($P$ = 2.76×10$^{-42}$). The imaging-based aging clock of kidney was specifically highly enriched in some metabolic pathways, such as sphingolipid metabolism ($P$ < 2.61×10$^{-68}$) and ether lipid metabolism ($P$ < 8.56×10$^{-5}$).

We also evaluated pleiotropic gene modules which were linked to multiple aging clocks. The enriched pathways of these gene modules allowed us to understand how cell biology perturbations have impact on molecular and structural aging in multiple organs. Two pleotropic gene modules involved multiple imaging-based aging clocks, which were artery and brain diffusion MRI (dMRI) as well as brain structural MRI (sMRI) and bone composition (**Fig. 3D**). The former pair highlighted the heart-brain connections, highly enriched in pathways such as ribosome biogenesis in eukaryotes ($P$ = 2.78×10$^{-69}$), which leads to reduced myocardial function as well as cognitive decline and synaptic damage. The gene modules shared by brain sMRI and bone composition aging clocks were enriched in several signaling pathways, such as wnt signaling pathway ($P$ = 1.90×10$^{-76}$), hippo signaling pathway ($P$ = 2.79×10$^{-47}$), signaling pathways regulating pluripotency of stem cells ($P$ = 7.32×10$^{-49}$), which play critical roles in tissue remodeling, regeneration, and homeostasis, including bone formation and neurogenesis. It is central to bone density regulation and structural brain maintenance, especially in aging. In addition, the shared gene module is also enriched in several neurodegenerative pathways, such as Alzheimer's disease ($P$ = $1.33\times10^{-43}$) as well as cancer pathways ($P$ = $7.63\times10^{-37}$).

We found five pleotropic gene modules that only involved protein aging clocks, which were (i) pancreas and adipose, (ii) pancreas and lung, (iii) liver and brain, (iv) liver and artery, and (v) adipose, liver and brain (**Fig. 3D**). Pathways such as other types of o-glycan biosynthesis were enriched in gene modules shared by pancreas and adipose ($P$ = $2.91\times10^{-2}$) as both tissues are highly involved in metabolic regulation and depend on glycosylation for their proper function and communication. SNARE interactions in vesicular transport were highly enriched in gene modules shared by pancreas and lung ($P$ = $2.76\times10^{-21}$). It is critical for vesicle-mediated transport, which is essential for processes like insulin secretion in the pancreas and surfactant release in the lung. Both pancreas and lung heavily rely on efficient vesicle trafficking to maintain their primary functions: insulin release from beta cells in the pancreas and surfactant release from alveolar cells in the lung. The enriched pathways in gene modules shared by liver and brain mainly reflect their interconnected roles in hormone metabolism and regulation, such as steroid hormone biosynthesis ($P$ = $1.23\times10^{-40}$) and ovarian steroidogenesis ($P$ = $8.12\times10^{-5}$). The gene module shared by the liver and heart was significantly enriched in complement and coagulation cascades ($P$ = $4.93\times10^{-14}$), likely due to the liver's central role in producing complement proteins and coagulation factors, which are vital for vascular integrity and immune regulation. The shared gene module between the brain, liver, and adipose tissue was enriched in pathways related to RNA regulation, including the spliceosome ($P$ = $1.02\times10^{-181}$), mRNA surveillance ($P$ = $2.18\times10^{-39}$), and RNA transport ($P$ = $2.56\times10^{-39}$). These pathways may highlight shared reliance on RNA regulation for neural function, metabolism, and stress responses.

**Additional details on physical activity traits**

The physical activity (PA) traits used in the phenotypic and genetic correlation were collected from UK Biobank (UKB) study. At the assessment center, participants were asked to answer touchscreen questions about the frequency and duration of various physical activities for the past 4 weeks (Category 100054). The self-reported PA types can be roughly characterized in terms of intensity into sedentary behaviors (e.g. driving, watching TV, using the computer), light PA (e.g. pruning, watering the lawn), moderate PA (e.g. cycling, walking for pleasure), vigorous PA (e.g. heavy lifting, aerobics), and other exercises to keep fit. From 2013 to 2015, the UKB study obtained consent from >100,000 participants to wear a wrist-worn triaxial accelerometer for seven consecutive days[1]. The acceleration signals captured at 100Hz were calibrated to local gravity, transformed to vector magnitudes by Euclidean norm, averaged in 5-second epochs, and stored under Field 90004. The advantage of device-based measurements compared to self-reported PA types is the capability to finely capture the PA intensity across the day. Therefore, stemming from the average vector magnitude values for every 5 seconds, we generated a series of accelerometry measurements covering the total activity level, activity in every 2-hour window of the day, active-sedentary transition patterns, and circadian rhythm proxies such as the activity level difference between active and inactive hours, referring to the workflow applied in Qi, et al. [2]. We only retained individuals with good calibration of the raw accelerometer data (Field 90016) and sufficient wear time (⩾3 days with accelerometer signals covering ⩾ 95% of the time). The derived measurements were rank-based inverse normal transformed, and measurements strongly correlated with others (absolute Pearson correlation coefficient > 0.8) were removed to reduce overlapping information. In addition to the PA intensity across the different times of the day, the duration of different PA types like self-reported data can also be inferred from the accelerometry signals using machine learning tools[3,4]. We downloaded data from the returned data 1942 and 2242 and Category 1020 for the proportion of time spent in some predicted PA types (e.g. walking, cycling, driving, overall light PA, overall moderate-to-vigorous PA, and overall sedentary behaviors). Finally, we kept 42 self-reported traits and 37 device-measured traits of PA, consisting of six categories: total PA, moderate-to-vigorous PA, light PA, sedentary behaviors, circadian rhythm, and other activity styles including walking pace, driving speed, active to sedentary transition patterns and other self-reported exercises.

**Additional details on sleep traits**

We used 34 sets of sleep genome-wide association studies (GWAS) summary statistics in the LDSC genetic correlation analysis with aging clocks. These included 5 GWAS on self-reported sleep duration[5-7] (including 1 short sleep duration and 1 long sleep duration), 5 accelerometer derived sleep duration[4,8,9] (including 1 short sleep duration and 1 long sleep duration), 2 on ease of getting up in the morning[6,7], 6 on chronotype[5-7,10,11], 2 on daytime napping[6,12], 5 on insomnia[6,7,13-15] , 4 on narcolepsy[6,7,16] (including 1 adjusted for BMI), and 5 on snoring[6,7,17,18](including 1 adjusted for BMI). The sleep traits examined in the phenotypic association study included several self-reported measures in the UKB study: (i) typical sleep duration ("About how many hours sleep do you get in every 24 hours? [including naps]", UKB Field 1160); (ii) ease of waking in the morning ("On an average day, how easy do you find getting up in the morning?", Field 1170; response scale from 1-4); (iii) chronotype ("Do you consider yourself to be a morning or evening person?", Field 1180); (iv) daytime napping frequency ("Do you have a nap during the day?", Field 1190); (v) insomnia symptoms ("Do you have trouble falling asleep or wake up during the night?", Field 1200); (vi) complaints of snoring from others ("Does your partner or a close relative or friend complain about your snoring?", Field 1210); and (vii) propensity to unintentionally fall asleep during the day ("How likely are you to doze off during daytime activities?", Field 1220). For all traits, individuals selecting "do not know" or "prefer not to answer" were excluded, and we used pre-coded responses provided by the UKB study.

**Further details on Mendelian randomization analysis**

To control for false positives, we applied Bonferroni correction for multiple testing within each disease category. The categories included: (1) brain disorders—comprising mental and behavioral disorders (F5), diseases of the nervous system (G6), and neurological endpoints; (2) cardiovascular diseases—including circulatory system diseases (I9_) and cardiometabolic endpoints; (3) abdominal organ and metabolic diseases—covering digestive system diseases (K11), genitourinary diseases (N14), respiratory system diseases (J10), endocrine, nutritional, and metabolic disorders (E4), as well as diabetes-related endpoints, asthma and related conditions, gastrointestinal comorbidities, and

COPD-related endpoints; (4) eye-related diseases—specifically diseases of the eye and adnexa (H7); (5) musculoskeletal disorders—diseases of the musculoskeletal system and connective tissue (M13); and (6) cancer—including neoplasms recorded in the cancer registry (ICD-O-3) and neoplasms from hospital discharge data (CD2). Additionally, we performed Bonferroni correction separately for each Mendelian Randomization (MR) method to avoid too stringent control. Additionally, we adjusted for multiple testing across both directions of MR jointly within each disease category—that is, both organ-to-disease and disease-to-organ directions were considered together during correction.

The primary Mendelian randomization (MR) results reported based on the inverse variance weighted (IVW) random effect method, which has been widely used. Results from other methods were included as sensitivity analyses. Specifically, to ensure the reliability of our results, we implemented several quality control procedures. We excluded causal estimates that relied on fewer than 4 genetic variants, as a larger number of genetic variants increases the statistical power of MR analysis[19,20]. We required reported causal pairs to be significant in IVW random effect model and at least three additional MR methods to avoid potential limitations or biases inherent to any single method. To further ensure the reliability of the results, we conducted several additional sensitivity tests. The MR-Egger intercept was used to check for directional horizontal pleiotropy. Higgins's $I^2$-test was used to evaluate heterogeneity among genetic instruments, as substantial heterogeneity might indicate violations of MR assumptions. As we performed bidirectional MR analysis, we also assessed the correctness of the causal direction using the mode estimate in GRAPPLE. If any causal pairs failed the above sensitivity tests (despite being significant in at least four MR methods), we required them to demonstrate significance in our robust MR methods. For example, if a selected causal pair failed in pleiotropy test, we require it to be significant in at least one MR method that account for pleiotropy effect. Similarly, if a selected pair fail the heterogeneity test, then we require it to be significant on at least one method that are robust to outliers. To minimize the instrumental bias, we require all selected causal pairs to be significant in at least one method that accounts for weak instrumental bias.

**MR power analysis**

We conducted power analysis[21] for both directions of two-sample MR to guide our assessment of required sample sizes. For clinical outcomes from the FinnGen study, the effective sample size was calculated using the following formula[22]:

$$n_{effective} = 4 \times n_{case} \times (n - n_{case})/\mathrm{n}$$

where n is the overall sample size. **Figure S4** illustrates the relationship between the number of cases, MR effect size, MR power, and variance explained by instrumental variables ($r^2$). For example, when aging clocks were used as exposure variables with $r^2$ = 0.02 and an MR effect size of 0.1 (supported empirically by our MR estimation results), statistical power exceeded 0.8 when the number of cases was greater than 10,000. Similarly, when FinnGen clinical endpoints were used as exposure variables with $r^2$ = 0.05 and an MR effect size of 0.05 (also supported empirically), statistical power exceeded 0.7 for cases greater than 10,000. Additionally, when the number of cases decreased to 6,000, the power did not drop dramatically, particularly when disease data were used as exposure variables. Under these conditions, the power remained above 0.5 for both MR directions. Furthermore, **Figure S3** summarizes the number of cases of our disease data, demonstrating that while we included certain diseases with smaller sample sizes, the majority of the disease (303/372) data in this study involved over 10,000 cases.

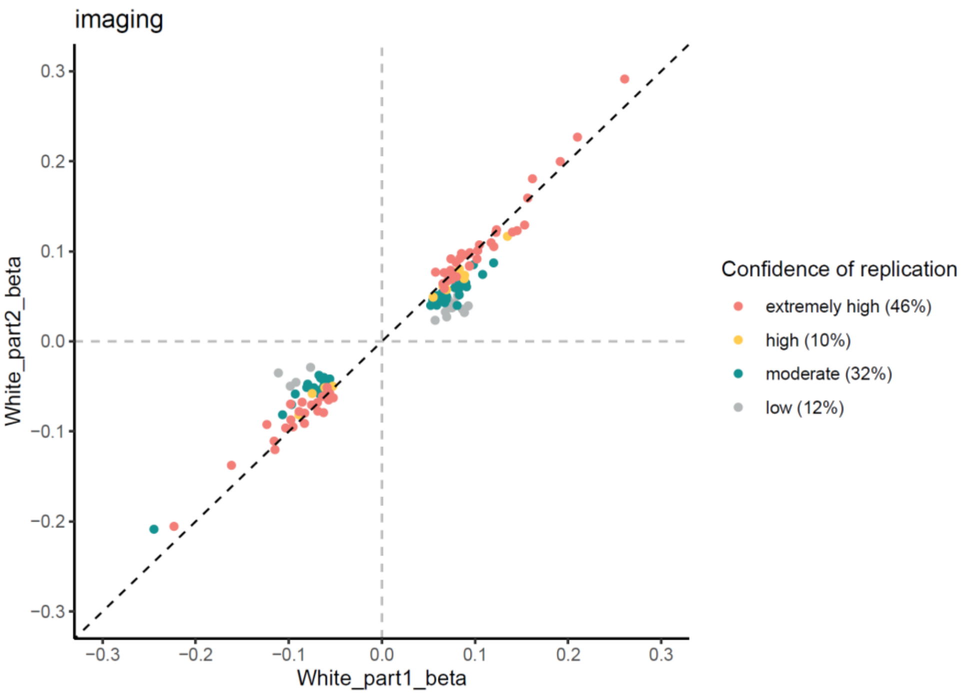


**Fig. S1. Consistency of genetic effect estimates between discovery and replication GWAS cohorts for imaging-based aging clocks.** The *x*-axis represents genetic effect estimates from the discovery cohort ($P < 2.08\times10^{-9}$) and the *y*-axis represents estimates from the replication cohort. Different colors represent the strength of replication evidence.

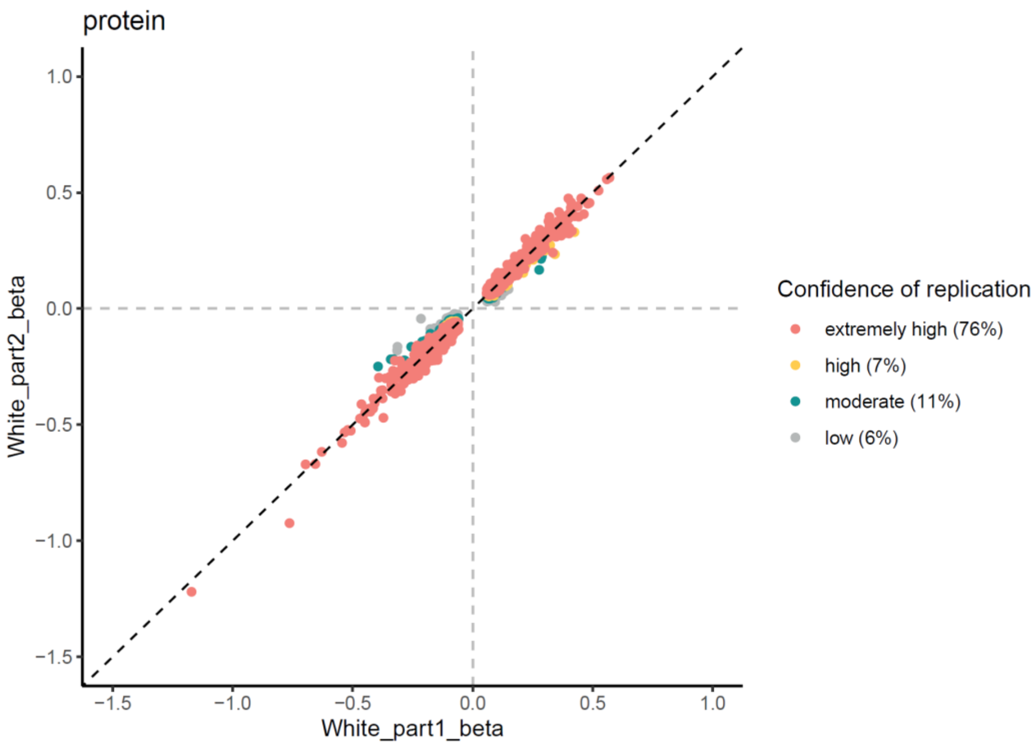


**Fig. S2. Consistency of genetic effect estimates between discovery and replication GWAS cohorts for protein-based aging clocks.** The *x*-axis represents genetic effect estimates from the discovery cohort ($P < 2.08\times10^{-9}$) and the *y*-axis represents estimates from the replication cohort. Different colors represent the strength of replication evidence.

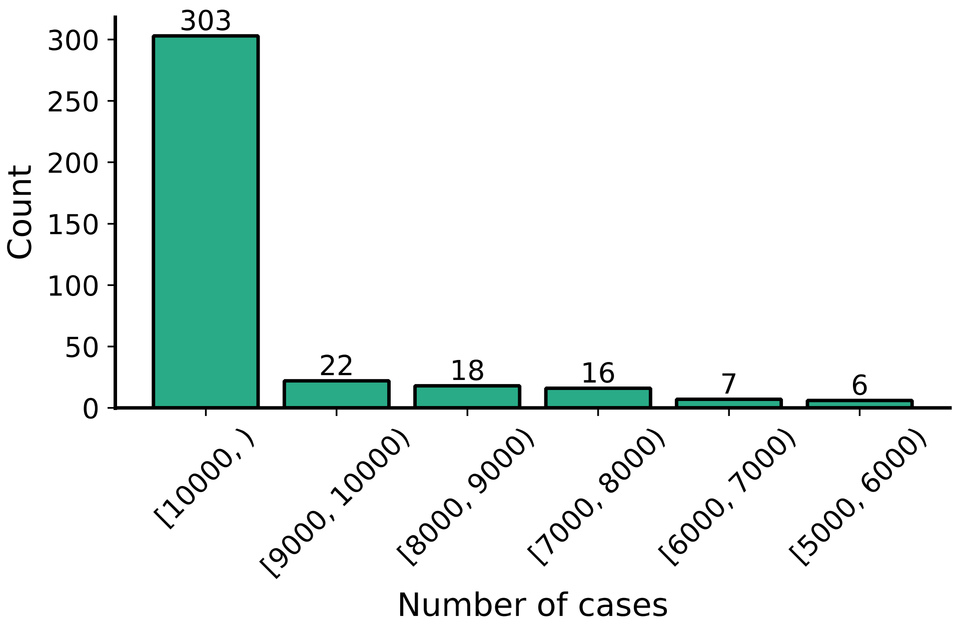


**Fig. S3. Distribution of number of cases for selected FinnGen clinical endpoints.** The *x*-axis represents case count intervals, and the *y*-axis indicates the number of endpoints falling within each interval. Details on the clinical endpoints are provided in **Table S33**.

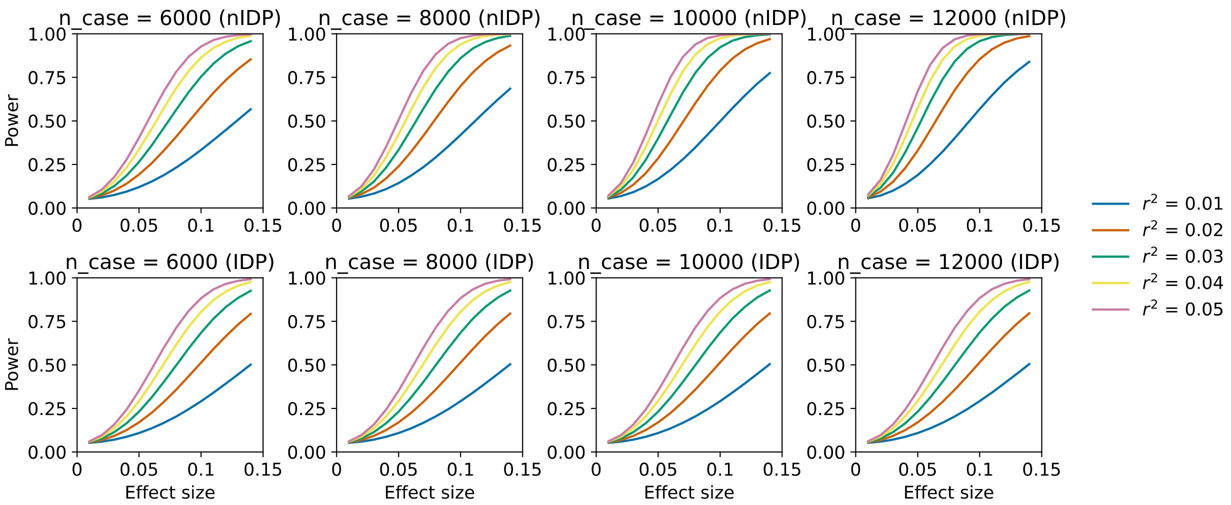


**Fig. S4. Mendelian randomization (MR) power analysis.** The relationship between MR effect size, number of cases, variance explained by instrumental variables ($r^2$), and statistical power is illustrated. Results are shown separately for both directions of Mendelian randomization: the upper panels depict aging clocks as exposures and FinnGen diseases as outcomes, while the lower panels show the reverse direction, with FinnGen clinical endpoints as exposures and aging clocks as outcomes.

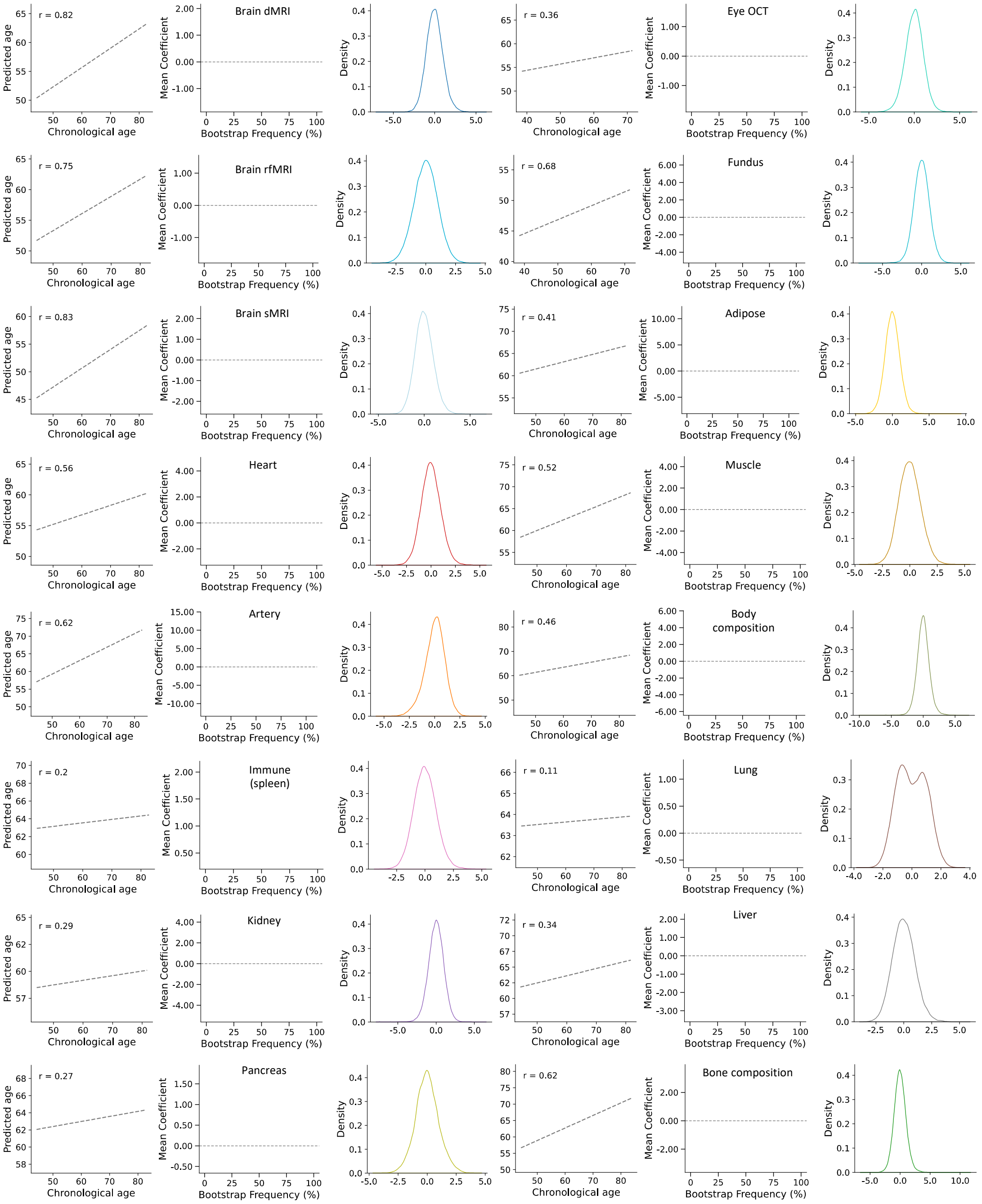


**Fig. S5. Prediction performance, feature importance, and distribution of imaging-based aging clocks.** For each imaging-based aging clock, three plots are presented: prediction accuracy between chronological and predicted age (left), feature importance (middle), and the distribution of aging clock scores (right). In the feature importance plot, bubble size and the *y*-axis indicate the average magnitude of the LASSO coefficients, while the *x*-axis represents the proportion of times each feature had a nonzero coefficient across 100 bootstrap iterations (**Methods**).

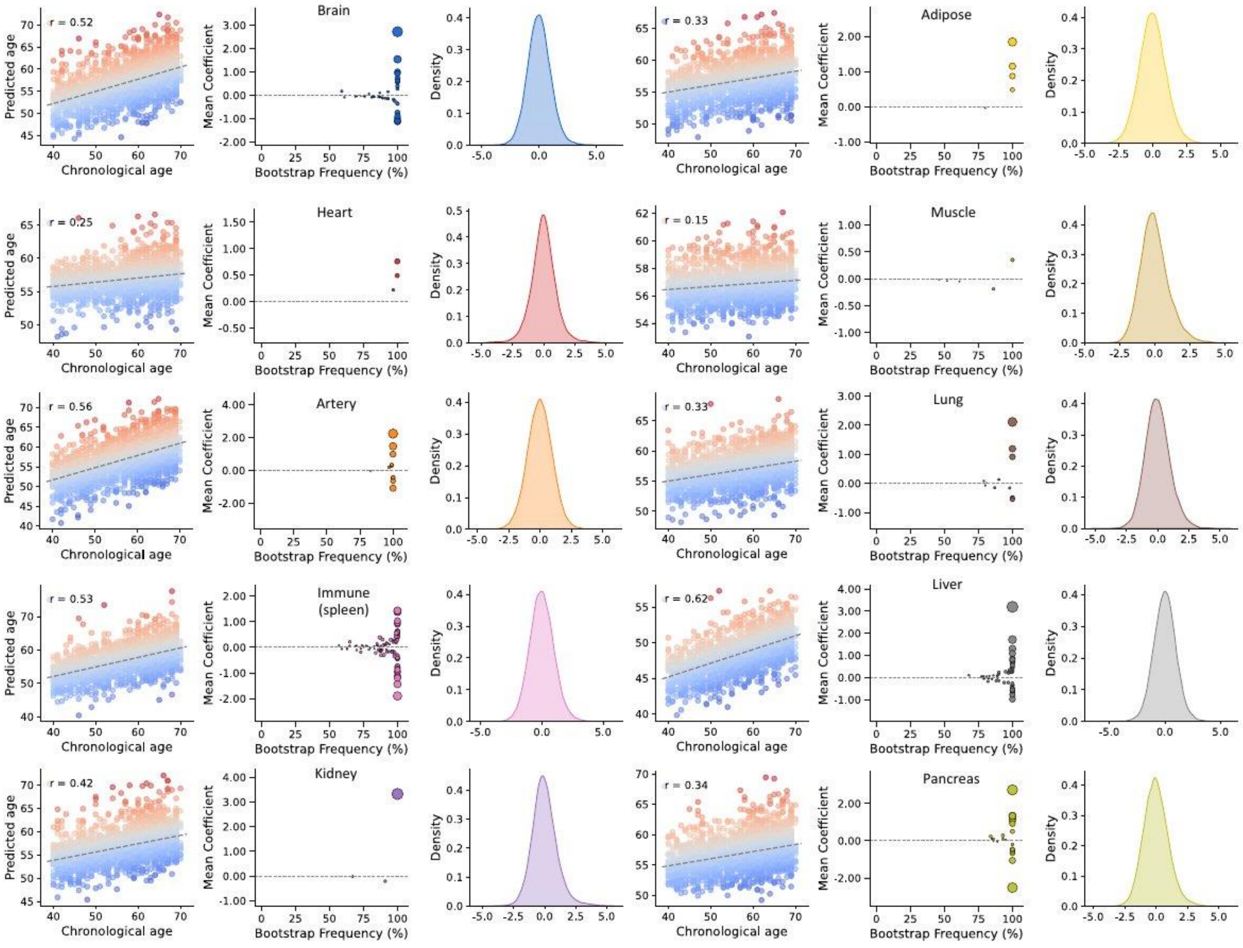


**Fig. S6. Prediction performance, feature importance, and distribution of protein-based aging clocks.** For each protein-based aging clock, three plots are presented: prediction accuracy between chronological and predicted age (left), feature importance (middle), and the distribution of aging clock scores (right). In the feature importance plot, bubble size and the *y*-axis indicate the average magnitude of the LASSO coefficients, while the *x*-axis represents the proportion of times each feature had a nonzero coefficient across 100 bootstrap iterations (**Methods**).
